# Land Use Barriers to Native and Western Food Security are Associated with Indigenous Maternal Death Occurrence and Changes to Nutritional Value of a Key Traditional Food

**DOI:** 10.1101/2025.07.24.25332149

**Authors:** CA Ricci, AE Orloff, M Wynne, L Pflueger, P Hallos, A Lopez, M Phelps, JM Postma, LE Hebert

**Affiliations:** Washington State University, College of Nursing; Washington State University, Institute for Research and Education to Advance Community Health (IREACH); Washington State University, Elson. S. Floyd College of Medicine; University of British Columbia, Institute of Resources, Environment and Sustainability; Spokane Tribal Network Tribal Food Sovereignty Initiative; Spokane Tribal Network Indigenous Birth Justice Initiative; Washington State University, Department of Animal Sciences; Bureau of Reclamation, Washington State, Yakima Basin; Washington State University, School of Biological Sciences

**Keywords:** pregnancy, food security, maternal health, maternal health disparities, Indigenous maternal health, AI/AN health disparities

## Abstract

For American Indian/Alaska Native (AI/AN) populations within the United States, nearly every pregnancy-related maternal mortality is preventable. Factors contributing to AI/AN pregnancy disparities are numerous, complex, and interacting, but there is recognition that diet and nutritional disparities have major roles. Many AI/AN individuals who are Western food insecure turn to traditional (Native) foods to augment general food security. Barriers to Native and Western food security exist, including those imposed by land use. We analyzed publicly available data to investigate relationships between agriculture (the number one land use purpose in the United States), Native and Western food securities, and the occurrence of AI/AN pregnancy-related maternal mortality. We additionally investigated agricultural associations with the nutritional status of a vital traditional food for AI/AN Peoples from Northern Pacific latitudes (coho Pacific salmon, *Oncorhynchus kisutch*). We demonstrate that relationships exist between food security, land use, and AI/AN pregnancy outcome. Relationships between land use and Native food security also indicate a need to consider the nutritional value of traditional foods consumed by pregnant AI/AN mothers.

## **1.** Introduction

American Indian/Alaska Native (AI/AN) health disparities are severe, pervasive, and extend to pregnancy. Pregnancy health disparities are of particular concern because the United States is in a maternal death crisis. Annual nationwide estimates of maternal deaths from pregnancy and its complications (i.e., pregnancy-related maternal death) reached >700 in 2019 (20.1 deaths per 100,000 live births)^1^. This was prior to the overturning of *Roe vs. Wade* (i.e., the 2023 *Dobbs Decision*), which has prevented many pregnant people from receiving life-saving preventative and/or emergency care^2^. State level analyses indicate that AI/AN women experienced the greatest increases in maternal death over recent decades, with median state maternal mortality ratios rising from 14.0 deaths per 100,000 births in 1999 to 49.2 in 2019^3^. Furthermore, retrospective analysis of AI/AN maternal deaths in the United States between 2017-2019 found that nearly every (>93%) pregnancy-related death was preventable^4^ (versus >84% in the general population^5^).

Factors contributing to AI/AN pregnancy disparities are numerous, complex, and interacting, but there is recognition that diet and nutrition disparities have major roles. For example, gestational diabetes is disproportionately prevalent among AI/AN populations (28.6 per 1,000 births in AI/AN populations compared to 8.7 per 1,000 births in non-Hispanic White populations in 2021^6^). Prevalences of several diet-related causes of death in AI/AN women of reproductive age (15-44 yrs) have also increased in the past two decades, including liver disease (+128.4%), cardiovascular disease (+55.3%), and diabetes mellitus (+213.0%)^7^. This has been concurrent with dietary shifts away from nutritionally healthy traditional foods and toward nutritionally deplete market foods^8^. Broad patterns of dietary shifts toward market foods are characterized by purchases of more affordable foods that tend to be highly processed and high in sugar, salt, and unhealthy fat, while also being low in beneficial nutrients^9^. These diets are most often a coping strategy for food insecurity, usually resulting from financial or geographic barriers to healthier options^10^.

Food security in academic literature typically applies a Western lens that follows the Food and Agriculture Organization of the United Nations (FAO) definition of, “when all people, at all times, have physical and economic access to sufficient and safe nutritious food that meets their dietary needs and food preferences for an active and healthy life.”^11^ Likewise, research in the United States often follows United States Department of Agriculture (USDA) recommendations for measuring food security according to whether individuals have “adequate” access to quality, variety, and desirability of foods^12^. However, many AI/AN and other Indigenous groups do not feel that these accurately reflect what food security encompasses for their Peoples^13^. The First Nations Development Institute, for example, alternatively defines Native food security as, “when American Indians, Alaska Natives, and Native Hawaiians, at all times, have access to an abundance of culturally relevant foods to meet their dietary needs and preferences for a healthy tribal community.”^14^ This definition shifts the benchmarks for food security from sufficiency to abundance, and from individual needs to whole community thriving. It is furthermore underscored by inherent and inextricable relationships between traditional foods, ancestral lands, Tribal sovereignty, and Indigenous Peoples. We utilize this definition in the analyses presented in the current study.

Many AI/AN individuals who are Western food insecure augment general food security with traditional foods, which can provide important nutrition^15^. For example, Pacific salmon species (a vital traditional food for AI/AN and other Indigenous Peoples from Northern Pacific latitudes) historically made up approximately 60% of the Spokane Tribe diet^15^ (situated in the eastern region of what we now call Washington state) and are high in nutrients that are known to support healthy pregnancy. A well-studied example is docosahexaenoic acid (DHA), an omega-3 polyunsaturated fatty acid, or PUFA, that is abundant in fish and is essential for neonate brain development^16^. Yet, challenges accessing traditional foods exist. Commonly cited barriers include environmental degradation (e.g., pollution, climate change) and the privatization and/or destruction of ancestral lands^17^. Economic factors such as cost of travel to ancestral hunting or harvesting areas (e.g., vehicle access, fuel, lodging, time off work) are also cited as barriers to accessing traditional foods^18^.

On the other hand, major facilitators of Native food security include social and community factors like food sharing and traditional knowledge sharing/transfer programs^18^. Some AI/AN leaders incorporate Indigenous and/or Tribal food sovereignty (I/TFS) principles into these programs as potential solutions to hunger and food insecurity in their communities. Indigenous Food Sovereignty can have different meanings between Indigenous groups but is generally regarded as a sacred responsibility held by Indigenous Peoples to participate in traditional practices that cultivate healthy and interdependent relationships with land, sea, air, soil, plants, and animals; and the cultural and traditional foods which are produced though these relationships are in turn viewed as sacred^19^. Similarly, Tribal food sovereignty has been described by cultural psychologist Dr. Melodi Wynne (Spokane Tribe of Indians) as the participation in connections among people, plants, elements, and all life above, on, in, and under the soil^20^.

We asked if land use had potential to influence AI/AN maternal health through food security and used agriculture as a representative land use variable (agriculture is the number one land use purpose within United States territories, comprising >1/3 of the total United States land area^21^). We first tested the nationwide distributions of agriculture for associations with Native and Western food security, and the occurrence of AI/AN pregnancy-related maternal death. Then as a case study, we investigated associations between nutritional characteristics of a traditional food (coho Pacific salmon, *Oncorhynchus kisutch*) sampled across a Pacific Northwest region (primarily Washington, United States). Nutritional quality was tested because changes to traditional foods that are available can have specific consequences for the Native food security of AI/AN Peoples including mothers. We acknowledge that poor pregnancy outcomes beyond maternal death occur (including long-term and severe maternal morbidity), and that poor pregnancy outcomes can additionally impact offspring health and development. AI/AN pregnancy-related maternal death was investigated here because of the United States maternal death crisis.

## **2.** Methods

### 2.1 Methodological overview

Associations between nationwide distributions of agriculture, Native and Western food security, and the occurrence of AI/AN pregnancy-related maternal death were tested at county level resolution. Data used were identified and compiled from multiple government and private foundation databases (CDC, USDA, Census Bureau, March of Dimes). We first parameterized county acreage dedicated to agriculture, impacts to Native food security, and levels of Western food security. We then identified associations between these characteristics and the occurrence of AI/AN pregnancy-related maternal deaths by comparing counties where a mortality was recorded (observation counties) to a random sample of counties where AI/AN pregnancy-related maternal death was not documented (non-observation counties).

Our case study examined nutritional characteristics of coho Pacific salmon (*Oncorhynchus kisutch*) that were directly measured from samples collected across Washington, United States (with one location in Idaho, United States). Land use and climate characteristics associated with coho Pacific salmon catch locations were obtained using GIS software. Coho Pacific salmon samples were obtained from localized fishers and businesses that were nearly exclusively AI/AN or AI/AN owned. Code for the analysis of coho Pacific salmon nutrition is available on GitHub (https://github.com/contessaricci/landuse-foodsecurity-AIANmatmort). Code for the analysis of AI/AN pregnancy-related maternal death is not published because this utilized restricted data. Additional details regarding methods are provided in the supplementary materials.

### 2.2 Data

Nationwide, AI/AN pregnancy-related maternal deaths were identified using the Centers for Disease Control and Prevention (CDC) Multiple Cause Mortality restricted use files^22^ for data years 2016-2019, and 2022. AI/AN pregnancy-related maternal deaths were identified as records from adult females aged 18 yr – 55 yr with i) the presence of an International Classification of Diseases, 10th Revision (ICD10) code indicating pregnancy, childbirth, and the puerperium (ICD10 O00-O9A) and/or an ICD10 code indicating obstetrical tetanus (A34) cited as a cause of death; and ii) any combination of AI/AN ancestry cited as the race. We balanced number of observation counties with non-observation counties by randomly selecting non-observation counties to equal the number of deaths recorded in a given state (final dataset: 122 observation counties; 131 non-observation counties). Records from Alaska were excluded due to differences in agricultural census data collection and availability protocols (n = 10).

Acreage of large agricultural operations at the county level was obtained using the 2017^23^ and 2022^24^ United States Department of Agriculture (USDA) agricultural census acreage estimates. The total agricultural acreage dedicated to operations ≥1,000 acres (sum of USDA farm size bins 1,000-1,999 acres, and >2,000 acres) was used as our definition of large agricultural operations.

Per county, variables for Native food security impacts, Western food insecurity experienced by the general population (generalized Western food insecurity), and Western food insecurity experienced specifically by AI/AN populations (AI/AN-specific Western food insecurity) were obtained using the 2019 USDA Food Access Research Atlas (FARA)^25^. Specific FARA variables used are listed in supplementary table 1. FARA variable values are aggregated census tract data to the county-level. Our variable “Native food security impacts” was a custom variable calculated by collapsing FARA variables that reflect factors documented in academic literature that can improve or hinder Native food security (see FARA variables and references listed in supplementary table 2).

County maternal care access rating was obtained from the March of Dimes Maternal Care Desert PeriStats query tool^26^. County land area (mi^2^) was obtained from the United States Census Bureau QuickFacts query tool^27^. State count of live births by mothers of AI/AN ancestry was obtained using the CDC Wide-ranging ONline Data for Epidemiologic Research (WONDER) query tool^28^.

### 2.3 GIS

GIS analysis was conducted using QGIS software^29^ (version 3.34) to obtain data associated with coho Pacific salmon (*Oncorhynchus kisutch*) catch locations. Distance from ocean was estimated as river miles (i.e., miles traveled along a river body) from a river’s estuary (i.e., where a river connects to the Pacific Ocean) to the catch location by tracing the most direct path along river tracts. Climate variables (i.e., temperature, precipitation), and agricultural intensity (% planted/cultivated) were quantified within a 15 mi radius buffer zone surrounding the coordinates of the catch location.

### 2.4 Coho Pacific salmon sample collection

Adult coho Pacific salmon migrating to spawning grounds during the 2022 fall run were collected for the current study. Samples were caught between October and November 2022. A total of 44 samples were obtained from 10 distinct water bodies: Chehalis River, Clearwater River, Dungeness Bay (Pacific Ocean, near shore), Nisqually River (Puget Sound), the outer coast (Pacific Ocean, open water), Peale Passage (Puget Sound), Port Orchard (Puget Sound), Salmon Bay Waterway (Puget Sound), Stillaguamish River, and Yakima River (figure 1). Catch locations spanned primarily across Washington, United States (n = 40), with samples from the Clearwater River collected in Idaho, United States (n = 4). GPS coordinates of catch locations were recorded for GIS analysis.

**Figure 1.**
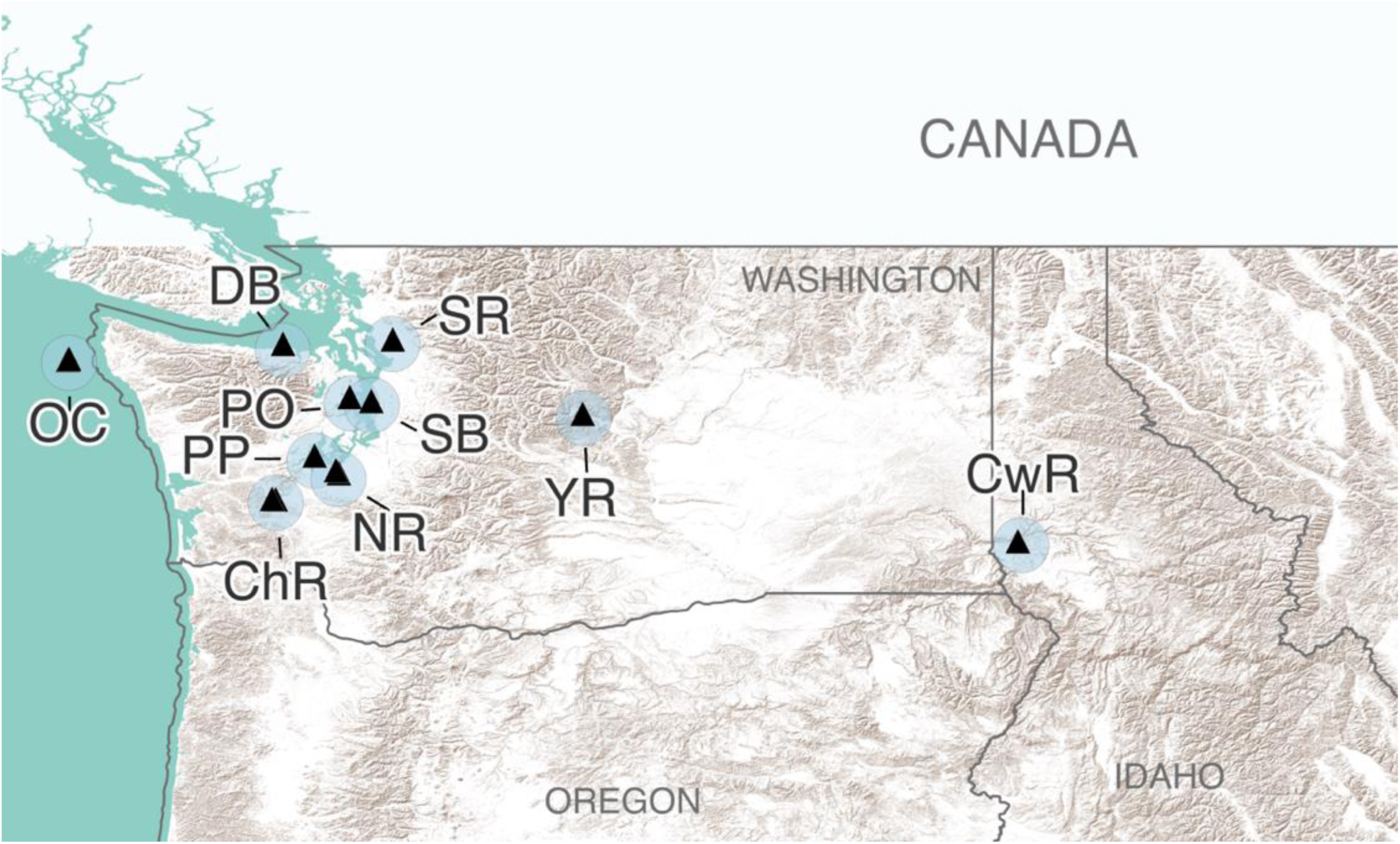
Catch locations for coho Pacific salmon samples. Exact catch locations are represented by black triangles; light blue circles surrounding catch locations are 15 mi buffer zones. ChR: Chehalis River; CwR: Clearwater River, DB: Dungeness Bay, NR: Nisqually River, OC: outer coast, PP: Peale Passage, PO: Port Orchard, SB: Salmon Bay Waterway, SR: Stillaguamish River, YR: Yakima River.

### 2.5 Coho Pacific salmon (Oncorhynchus kisutch) nutritional analysis

Nutritional analysis was performed by Palouse Environmental Services (Pullman, WA, United States) on muscle tissues collected from coho Pacific salmon samples. Nutritional analysis included a proximate nutrition analysis (ash, moisture, crude protein, crude fat), a fatty acid methyl esters (FAMEs) profile (35 fatty acids measured), and total mercury (THg).

### 2.6 Statistical analysis

All statistical analyses were conducted using R statistical software (version 4.4.1)^30^. Multivariate outlier detection for both analyses was carried out using median absolute deviation (MAD)^31^. MAD for analysis of AI/AN pregnancy-related maternal death in relation to food security and agriculture used a cutoff at 4 standard deviations and was based on: outcome (observation, non-observation), large agricultural operations acreage, county land area, state count of live births by mothers of AI/AN ancestry, county urbanization, county averaged median family income, county AI/AN population count, generalized Western food insecurity, AI/AN-specific Western food insecurity, and vehicle insecurity. MAD for coho Pacific salmon nutritional analysis used a cutoff at 3 standard deviations for outlier determination and was based on: fish weight (kg), distance from ocean, urbanization, agricultural intensity, climate (temperature, precipitation), and proximate nutrition analysis. FAMEs profile was not included because FAMEs variables are a proportion of crude fat. Fish age was also not included because a large proportion of fish (n = 30, 68.18% of samples) were aged the same (3 yrs).

Model weighting used Inverse Probability of the Treatment Weighting (IPTW)^32^. IPTW for analysis of AI/AN pregnancy-related maternal death represented probability of mortality and was based on: year of mortality occurrence, county rating for access to maternal care, county land area, and state count of live births from mothers of AI/AN ancestry. Coho Pacific salmon nutritional analysis alternatively used IPTW for multiple treatments^33^ and represented probability of belonging to true water body of origin. Coho Pacific salmon IPTW was based on sex, weight, distance from ocean, buffer zone urbanization, and hatchery marking.

The relationship between AI/AN pregnancy-related maternal death, Native and Western food security, and acreage dedicated to large agricultural operations was determined using model comparison of generalized linear models with binary distributions. Our “Native food security impacts” variable was calculated using the PC1 eigenvalues for robust-scaled FARA variables that were collapsed by principal component analysis (PCA, supplementary figure 1). A total of 11 models were compared (supplementary table 3) and a Bonferroni multiple corrections significance threshold of p ≤ 0.004 was enforced. Model performance was based on goodness of fit. Goodness of fit variables considered were Tjur’s R^2^ for binary outcomes, root mean squared error (RMSE), and % correct predicted (PCP).

Coho Pacific salmon nutrition responses to agricultural intensity and climate were determined by multivariate linear modeling using nutritional analysis outcomes in 3 separate models (proximate analysis, FAMEs profile, and THg) and enforced a Bonferroni multiple comparisons significance threshold of p ≤ 0.017. We treated the proximate analysis and FAMEs profile as a univariate response variable by collapsing proximate nutrition variables and fatty acids (respectively) by PCA and used the PC1 eigenvalues as the nutrition summary. Likewise, we calculated a “climate” variable using PCA due to a high correlation between temperature and precipitation (supplementary figure 3). The PC1 eigenvalue was used as the “climate” value.

## 3. Results

### 3.1 Occurrence of AI/AN pregnancy-related maternal death can be partly explained by relationships between acreage dedicated to large agricultural operations, impacts to Native food security, and generalized Western food insecurity

Of the 11 generalized linear models compared, the model using county-level acreage dedicated to large agricultural operations, Native food security impacts, and generalized Western food insecurity explained the occurrence of AI/AN pregnancy-related maternal death best (Tjur’s R^2^ = 0.292, RMSE = 0.429, PCP = 0.646, table 1). Generally, Tjur’s R^2^ for univariate models did not exceed 0.02, with exception of Native food security impacts, which nearly reached 0.2 (table 1). Also, county-level maternal care access did not explain AI/AN pregnancy-related maternal death occurrence. The majority of counties rated highest for maternal care access (∼60%) were associated with occurrence of AI/AN pregnancy-related maternal death, while the majority of counties classified as maternal care deserts (∼79%) were those where AI/AN pregnancy-related maternal death was not observed (figure 2A). Standalone variables included in the most explanatory model were also not individually informative to occurrence of AI/AN pregnancy-related maternal death (figure 2B-D).

**Table 1.** Model comparisons. Model goodness of fit comparisons for county-level factors associated with AI/AN pregnancy-related maternal death occurrence. Models arranged in descending order from best fit to worst fit.

| Model | Variables | Tjur's R<sup>2</sup> | RMSE | PCP |
| --- | --- | --- | --- | --- |
| J | Native food, Western food (gen), ag acreage | 0.292 | 0.429 | 0.646 |
| K | Native food, Western food (AI/AN), ag acreage | 0.250 | 0.436 | 0.625 |
| E | Native food, Western food (gen) | 0.226 | 0.438 | 0.613 |
| F | Native food, Western food (AI/AN) | 0.195 | 0.441 | 0.597 |
| C | Native food | 0.194 | 0.441 | 0.597 |
| I | Native food, ag acreage | 0.190 | 0.445 | 0.595 |
| H | Western food (AI/AN), ag acreage | 0.057 | 0.485 | 0.528 |
| G | Western food (gen), ag acreage | 0.038 | 0.494 | 0.518 |
| A | Western food (gen) | 0.013 | 0.498 | 0.506 |
| B | Western food (AI/AN) | 0.008 | 0.498 | 0.504 |
| D | Ag acreage | 0.001 | 0.500 | 0.500 |

*Native food: county Native food security impacts; Western food (gen): county generalized Western food insecurity; ag acreage: county acreage dedicated to large agricultural operations; RMSE: root mean squared error; PCP: % correct predicted*

**Figure 2.**
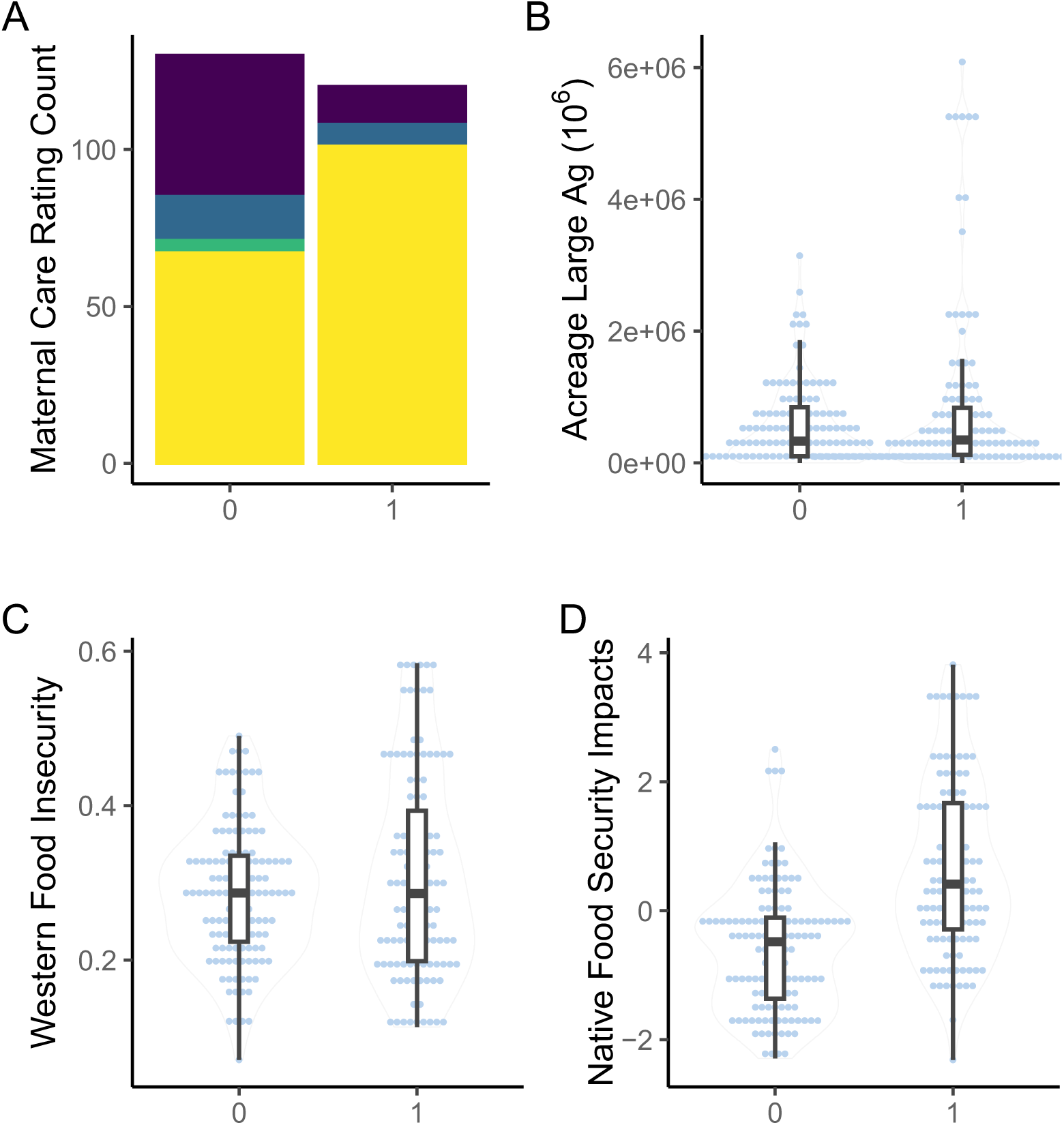
Univariate comparisons. Panel plot of univariate distributions between observation counties (1) and non-observation counties (0). Box plots superimposed on violine plots (B-D) represent distribution of observation and non-observation counties sampled. (A) stacked bar plot of county maternal care access rating. Height of bar represents count of counties included in analysis that are rated respectively. Yellow: highest rating of maternal care access; green: moderate maternal care access; blue: low maternal care access; purple: maternal care desert. (B) volin plot of acreage dedicated to large agricultural operations. (C) violin plot of generalized Western food insecurity; (D) violin plot of Native food security impacts.

Variables included in the most explanatory model reached Bonferroni multiple comparisons significance for Native food security impacts (p < 0.001, table 2), generalized Western food insecurity (p = 0.004, table 2), the interaction between Native food security impacts and agricultural acreage dedicated to large agricultural operations (p = 0.001, table 2), and the interaction between all 3 variables (p = 0.002, table 2). AI/AN pregnancy-related maternal death occurrence was primarily driven by Native food security impacts, followed by generalized Western food insecurity, and then followed by the interaction between all 3 variables (figure 3). AI/AN pregnancy-related maternal death occurrence separated above and below the IQR of county-level Native food security impacts. Counties with very high impacts to Native food security (>3^rd^ quartile, 0.672) were primarily those where mortality was observed, and counties with very low impacts to Native food security (<1^st^ quartile, -0.994) were primarily those where mortality was not observed (figure 4). Patterns of generalized Western food insecurity were not associated with mortality at lower agricultural acreages, however, generalized Western food insecurity increased concurrent with county agricultural acreage (figure 4). At very high county agricultural acreages (>3^rd^ quartile, >845,289 acres, 51 counties), counties with very low impacts to Native food security comprised approximately 1/5 of counties (21.569%), counties with high Western food security (>median, 0.287) comprised approximately 1/3 of counties (33.333%), and counties with both very low impacts to Native food security and high Western food security comprised approximately 1/10 of counties (11.764%)(supplementary table 4). In all categories, higher food security was associated with counties that did not observe AI/AN pregnancy-related maternal death. The greatest disparities were present in counties with very low impacts to Native food security and in counties with both very low impacts to Native food security and high Western food security (supplementary table 4).

**Table 2.** Model J. Results for the model most explanatory to AI/AN pregnancy-related maternal death occurrence. Statistically significant effects were subject to a Bonferroni multiple comparisons threshold of p-value ≤ 0.004.

|  | Est | SE | t val | p-val |
| --- | --- | --- | --- | --- |
| **Native food** | **1.144** | **0.213** | **5.382** | **1.720<sup>e-7</sup>** |
| **Western food (gen)** | **1.018** | **0.262** | **3.883** | **1.330<sup>e-4</sup>** |
| **Native food X ag acreage** | **-0.594** | **0.183** | **-3.251** | **0.001** |
| **Native food X Western food (gen) X ag acreage** | **0.780** | **0.249** | **3.132** | **0.002** |
| Native food X Western food (gen) | -0.451 | 0.279 | -1.619 | 0.107 |
| Western food (gen) X ag acreage | -0.443 | 0.307 | -1.443 | 0.150 |
| Ag acreage | 0.065 | 0.230 | 0.281 | 0.779 |

*Native food: county Native food security impacts; Western food (gen): county generalized Western food insecurity; ag acreage: county acreage dedicated to large agricultural operations; Est: estimate; SE: standard error; ag: agriculture; null deviance: 693.97 on 249 degrees of freedom; residual deviance: 517.80 on 242 degrees of freedom; bolded p-values represent predictors meeting Bonferroni significance threshold*

**Figure 3.**
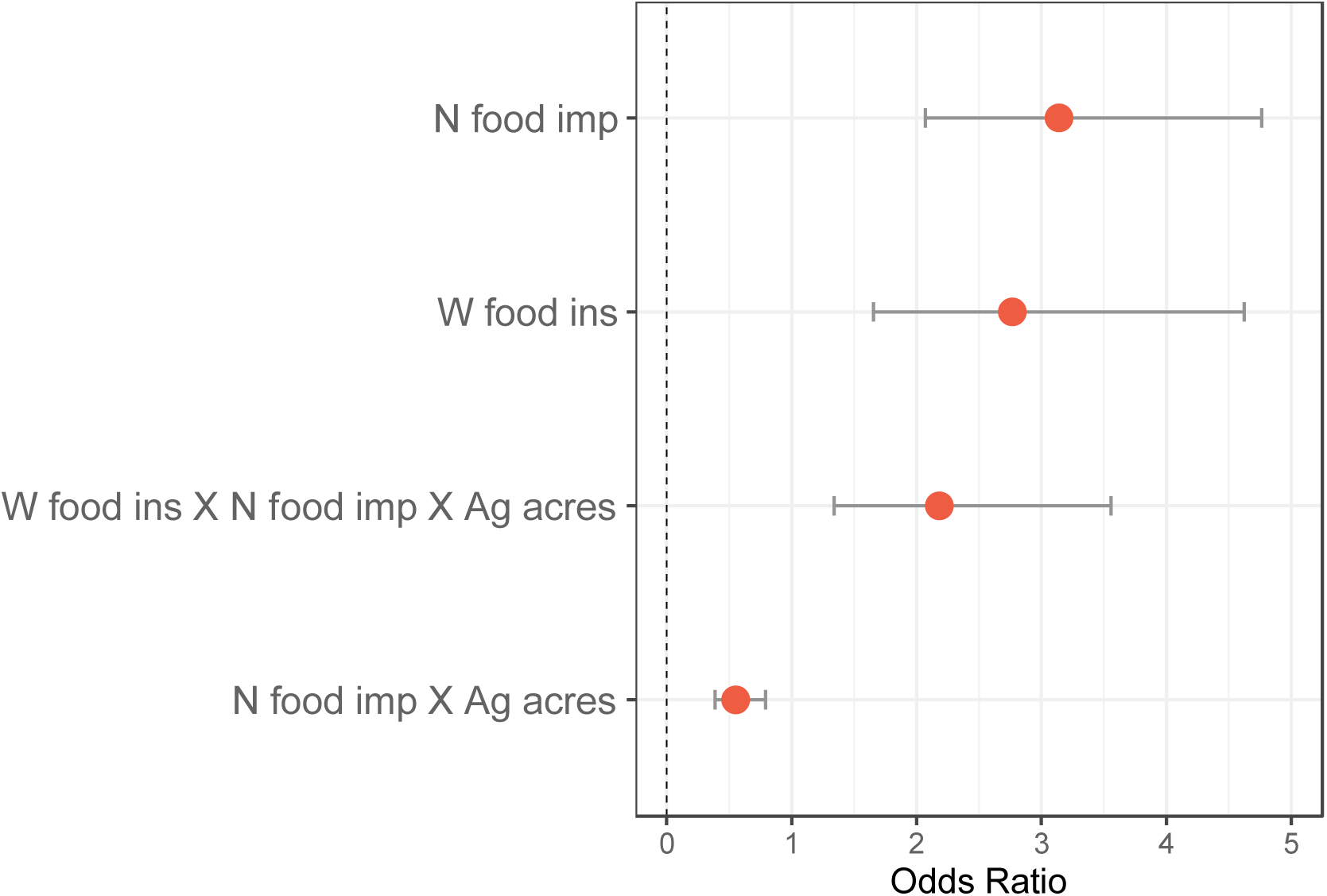
Model J odds ratio plot. Odds ratio plot for effect size of variables included in most informative model that contribute to AI/AN pregnancy-related maternal death occurrence. Variables not reaching Bonferroni multiple comparisons significance threshold (p > 0.004) are not shown. N food imp: county Native food security impacts; W food ins: county generalized Western food insecurity; Ag acres: county acreage dedicated to large agricultural operations.

**Figure 4.**
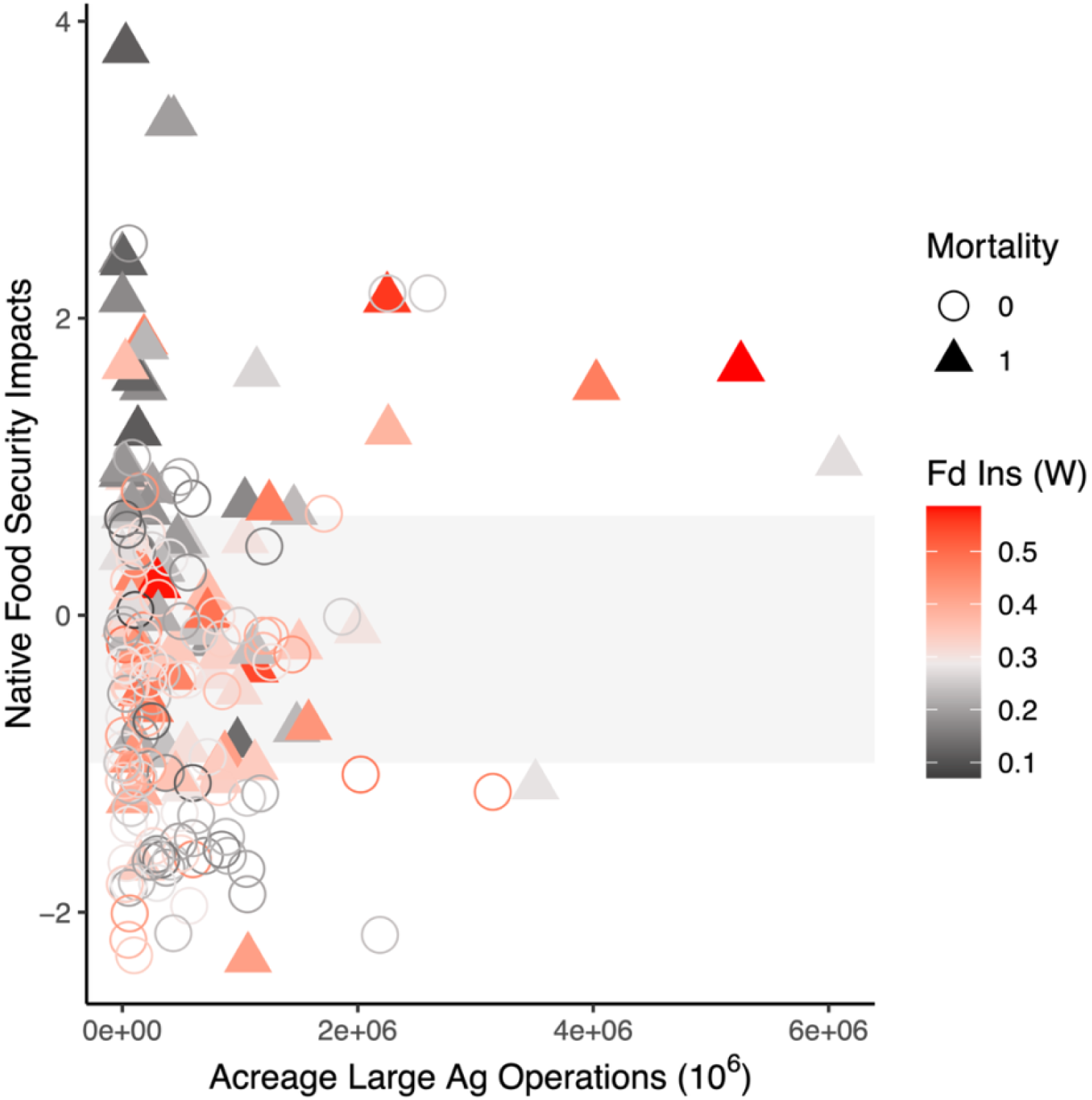
AI/AN pregnancy-related maternal death occurrence in relation to county acreage dedicated to large agricultural operations, county Native food security impacts, and county generalized Western food insecurity. Filled triangles represent observation counties; open circles represent non-observation counties. Symbol color corresponds to county generalized Western food insecurity with midpoint between shades set at median level of generalized Western food insecurity across all counties included in analysis. Shaded horizontal bar represents the IQR for Native food security impacts (bottom: 1^st^ quartile, top: 3^rd^ quartile) across all counties included in analysis. Native food security impact values above IQR represent very high impacts to Native food security; values below IQR represent very low impacts to Native food security.

### 3.2 Agricultural intensity and climate (precipitation, temperature) are associated with changes to the nutritional status of coho Pacific salmon

THg was significantly explained by climate, agricultural intensity, and their interaction (table 3). FAMEs profile was significantly explained by agricultural intensity (table 4). A nearly significant relationship between proximate nutrition and agricultural intensity was observed but did not reach the Bonferroni multiple comparisons threshold (table 5). Levels of THg increased with agricultural intensity (figure 5). Differences in interactions between climate variables (precipitation, temperature) and agricultural intensity were observed, specifically, THg levels increased more in relation to agricultural intensity when fish were in climates characterized by higher precipitation and cooler temperatures. However, this study was not powered to test the statistical significance of this relationship.

**Table 3.** Total mercury (THg). Results for THg associations with agricultural intensity and climate. Statistically significant effects were subject to a Bonferroni multiple comparisons significance threshold p-value ≤ 0.017

|  | **Est** | **SE** | **t val** | **p-val** |
| --- | --- | --- | --- | --- |
| % agriculture | **0.008** | **0.001** | **7.450** | **7.180<sup>e-9</sup>** |
| climate | **-0.047** | **0.012** | **-3.829** | **4.800<sup>e-4</sup>** |
| % agriculture x climate | **0.010** | **0.002** | **5.703** | **1.590<sup>e-6</sup>** |

*Multiple R-squared: 0.6987; adjusted R-squared: 0.6743; model p-value: 9.608e<sup>-10</sup>; bolded p-values represent predictors meeting Bonferroni significance threshold*

**Table 4.** FAMEs profile. Results for FAMEs profile associations with agricultural intensity and climate. Statistically significant effects were subject to a Bonferroni multiple comparisons significance threshold p-value ≤ 0.017

|  | **Est** | **SE** | **t val** | **p-val** |
| --- | --- | --- | --- | --- |
| % agriculture | **0.223** | **0.074** | **3.021** | **0.005** |
| climate | 0.347 | 0.838 | -0.414 | 0.681 |
| % agriculture x climate | 0.132 | 0.123 | 1.083 | 0.286 |

*Multiple R-squared: 0.520; adjusted R-squared: 0.4811 model p-value: 4.623<sup>e-6</sup>; bolded p-values represent predictors meeting Bonferroni significance threshold*

**Table 5.** Proximate nutrition profile. Results for proximate nutrition profile associations with agricultural intensity and climate. Statistically significant effects were subject to a Bonferroni multiple comparisons significance threshold p-value ≤ 0.017

|  | **Est** | **SE** | **t val** | **p-val** |
| --- | --- | --- | --- | --- |
| % agriculture | 0.139 | 0.059 | 2.359 | 0.024 |
| climate | -0.503 | 0.672 | -0.749 | 0.459 |
| % agriculture x climate | 0.144 | 0.098 | 1.460 | 0.152 |

*Multiple R-squared: 0.2319; adjusted R-squared: 0.1696; model p-value: 0.01952*

**Figure 5.**
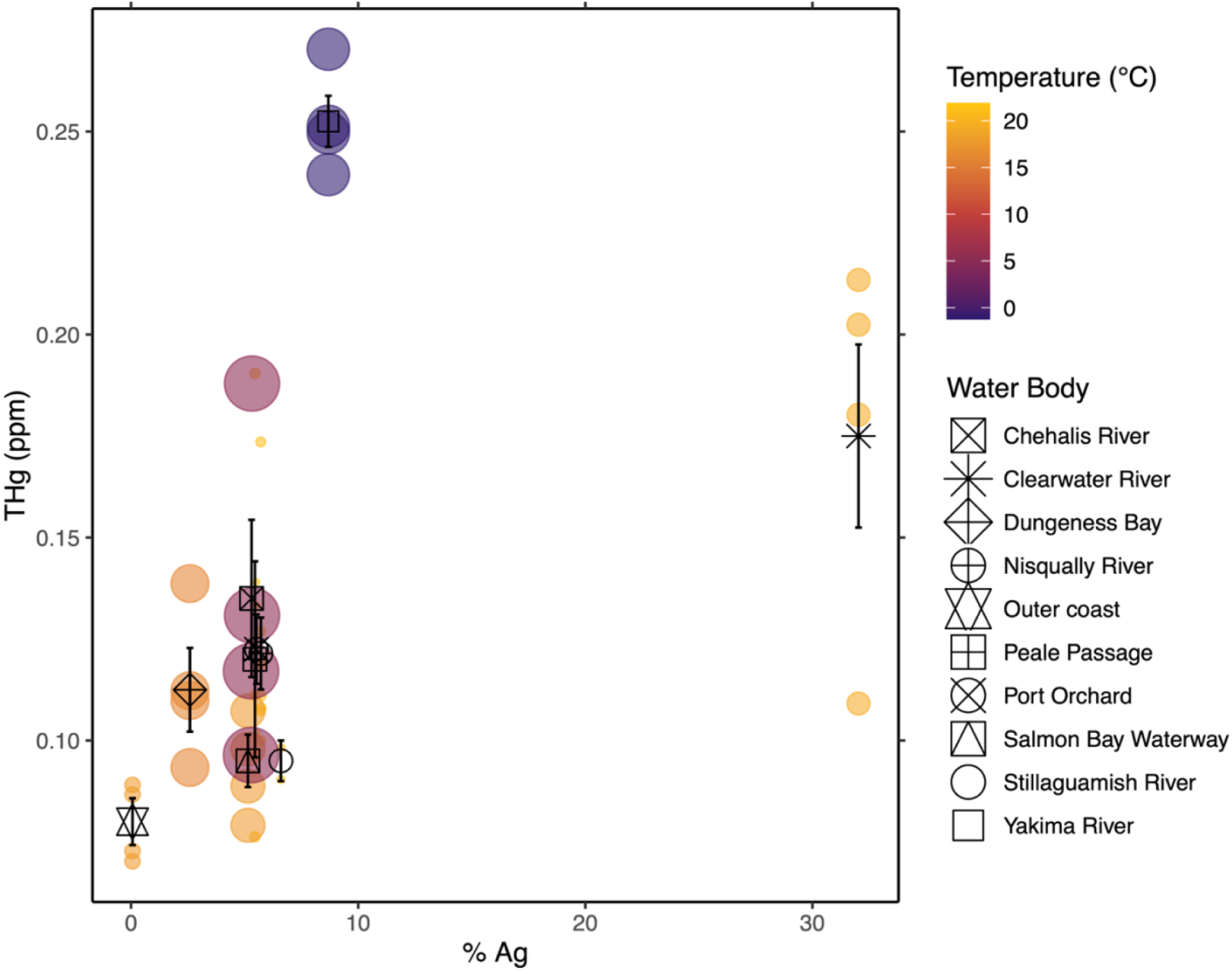
Total mercury. Scatterplot showing relationship between total mercury (THg) and agricultural intensity (% Ag), mediated by temperature and precipitation (climate). Temperature is mean temperature of previous month measured at catch location (°C; min:-1.22 C, max: 21.86 C). Precipitation is mean precipitation of previous month measured at catch location (inches; min: 0.08 in, max: 11.07 in). Colored bubbles represent individual samples. Color of bubbles represents temperature; size of bubbles represents precipitation. Black symbols represent mean total mercury of samples grouped by water body of origin; vertical error bars through black symbols represent standard error of total mercury.

FAMEs profile relationships were investigated by ordination (redundancy analysis, RDA). FAMEs most responsive to agricultural intensity were those at or exceeding the 3^rd^ quartile of r^2^ values for ordination goodness of fit (r^2^ ≥ 0.44). The most responsive FAMEs (n = 7) were docosahexaenoic acid (DHA), eicosadienoic acid (EA), gamma-linolenic acid (GLA), myristic acid (MA), pentadecanoic acid (PDA), palmitic acid (PA), and palmitoleic acid (POA). 6 of the 7 FAMEs (DHA, EA, GLA, MA, PDA, POA) have known or putative health benefits and 1 FAME (PA) imparts health risks in a maternal lipid-imbalance context (supplementary table 7). With exception of DHA, all eigenvectors for the most responsive FAMEs (EA, GLA, MA, PDA, PA) were primarily opposite to agricultural intensity increases along the PC1 axis, which explained most of the variation observed across FAMEs profiles (61.67%, figure 6). The DHA eigenvector was approximately perpendicular to the 6 other FAMEs along the PC2 axis (15.37% variation explained, figure 6), indicating that other factors in addition to agricultural intensity significantly influenced DHA levels and that there may be a tradeoff between levels of DHA and the levels of other 6 FAMEs. Fish collected from the Clearwater River were associated with the highest agricultural intensity and had FAMEs profiles characterized by lower levels of EA, GLA, MA, PDA, and PA. Fish collected from the Yakima River, the Chehalis River, Dungeness Bay, and the outer coast, as well as select fish from other water bodies (Port Orchard, Clearwater River), were higher in DHA and lower in other 6 FAMEs, and with levels of other 6 FAMEs increasing in association with decreases in agricultural intensity. Profiles primarily characterized by higher levels of EA, GLA, MA, PDA, and PA were associated with fish collected from Puget Sound areas.

**Figure 6.**
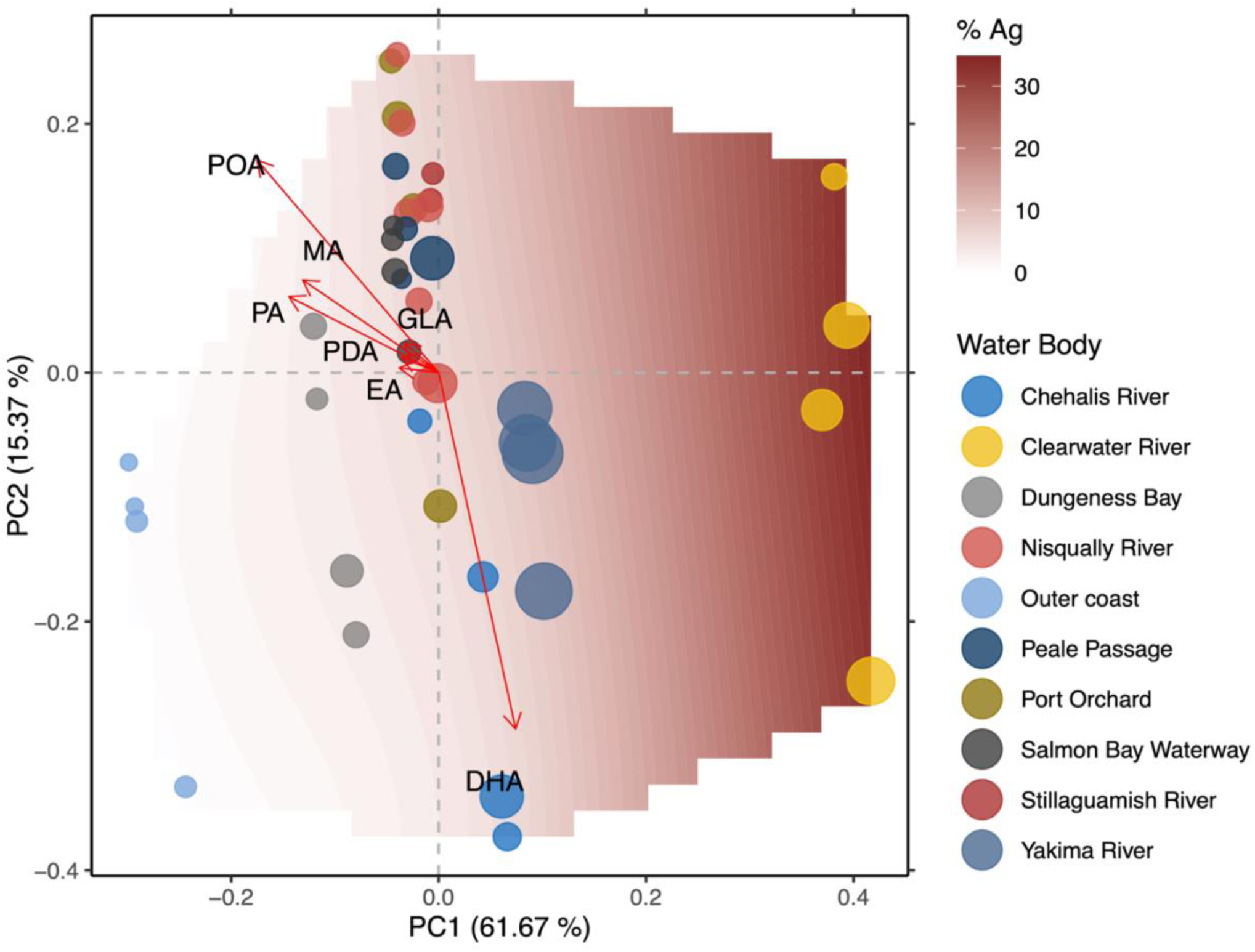
FAMEs. Ordinal plot showing relationship between sample-weighted fatty-acid methyl esters (FAMEs) profiles and agricultural intensity (% agricultural land use). Contour surface represents agricultural intensity, with white and blue shades representing low levels of agricultural intensity that transition to deeper shades of burgundy with greater levels of agricultural intensity. Colored bubbles represent individual samples. Color of bubbles correspond to water body of origin, size of bubbles contextualizes relative levels of total mercury measured in samples (parts per million; min: 0.07 ppm, max: 0.27 ppm). Vectors represent magnitude and directional response of FAMEs with significantly and strongly responding to agricultural intensity (r > 0.88, p < 0.05). Specific FAMEs corresponding to vector arrows are labeled in black. DHA: docosahexaenoic Acid; EA: eicosadienoic acid; GLA: gamma-linolenic acid; MA: myristic acid; PDA: pentadecanoic acid; PA: palmitic acid; POA: palmitoleic acid

## 4. Discussion

Maternal diet and nutrition at periconception and during pregnancy are leading modifiable risk factors in the development of poor pregnancy outcomes, including for AI/AN mothers^34^. Also, the inability to attain optimal nutrition to support pregnancy is related to structural systems that determine vital components of human livelihood (i.e., social and structural determinants of health such as income, vehicle access, or proximity to medical services; and cultural determinants of health such as land, intellect, spirit, family, community). AI/AN Peoples experience disproportionate rates of Western food insecurity^35^ and poor pregnancy and maternal health outcomes^4^, concurrently. When experiencing Western food insecurity, some individuals augment diets with traditional foods^36^. We considered both Native and Western food security here, which are infrequently explored together quantitatively in academic literature and especially in the context of AI/AN pregnancy and maternal health disparities.

We show that very high impacts to Native food security and high levels of Western food insecurity experienced by a county’s general population are associated with greater occurrence of AI/AN pregnancy-related maternal death, and that agricultural land use influences this association. At very high agricultural acreages, both Native and Western food secure counties are rare. Furthermore, Native food secure counties are especially skewed toward those that did not observe any AI/AN pregnancy-related maternal death. These data corroborate traditional knowledge systems that assert food security, and Native food security in particular, as protective in AI/AN pregnancy health^37^. These data also suggest that the relationship between maternal diet and AI/AN pregnancy-related maternal death is exaggerated when mothers are living in association with high agricultural acreages.

Additionally, agricultural intensity and climate were dynamic predictors of the nutritional status of a key traditional food, coho Pacific salmon (*Oncorhynchus kisutch*). Quantities of the beneficial fatty acids eicosadienoic acid (EA), gamma-linolenic acid (GLA), myristic acid (MA), pentadecanoic acid (PDA), and palmitoleic acid (POA) decreased in response to agricultural intensity. Quantities of docosahexaenoic acid (DHA) also decreased in response to agricultural intensity but patterns of geographic distributions of DHA quantities in coho Pacific salmon in relation to the other most responsive fats suggest there may be tradeoffs between DHA content and the synthesis of other beneficial fats. Agricultural activity in our sampling region increases with distance from the Pacific Ocean. Pacific salmon cease eating upon commencing spawning migrations and so the distance Pacific salmon travel upriver will influence muscle tissue composition and energy reserves^38^. Including this confounder as a weighting variable in our analyses allowed examination of nutritional relationships with agriculture while enforcing an artificially equal distribution of distance from ocean across samples^32^. DHA, EA, GLA, MA, PDA, and POA are essential for various functions that support neonate/infant growth and development (among others) and also have roles in oocyte maturation and maintaining maternal health during pregnancy. One fatty acid, palmitic acid (PA), has been implicated as a risk to pregnancy health in lipid imbalance contexts such as that in maternal obesity or when maternal diets are high in unhealthy fats. On the other hand, PA was associated with decreased risk of asthma development in children born to mothers with healthy diets, is a necessary fatty acid for cellular membranes, and endogenously synthesized PA is a major component of both fetal adipose tissues and fatty acids in maternal milk in normal pregnancy (see references in supplementary table 7).

Total mercury increased in response to agricultural intensity and climates characterized by cooler temperatures with higher precipitation. The relationship between total mercury levels with respect to agricultural intensity also appears to magnify with increased precipitation, although our study was not powered to statistically test this association. We did not determine sources of mercury present in coho Pacific salmon, but dietary mercury is understood to be the dominant route of exposure in fish^39^. However, our results indicate environmental exposure rather than dietary exposure because mercury levels increased as distance from ocean (i.e., assumed last food intake) increased. Aqueous mercury exposure occurs^39^ but the toxicokinetics of this route have not been studied in adult Pacific salmon (to our knowledge). In mosquito fish (*Gambusia affinis*) and redear sunfish (*Lepomis microlophus*), aqueous mercury is retained at rates higher than mercury removal rates, and the majority of dietary mercury is removed after 48 h while the majority of aqueous mercury is retained^39^. Nevertheless, mercury retention and removal rates vary across species and are significantly influenced by multiple factors including water salinity, fish weight, and form of mercury^40^. Relevant to the current study, it has been hypothesized that cold and wet climates in northern Europe promote the build-up of organic materials, including mercury, in soils associated with agriculture and adjacent lands^41^. In this context, our data implicate settings associated with agricultural land as a constituent source of environmental mercury that migrating salmon are exposed to, with precipitation possibly functioning to enhance salmon exposure levels via increased transportation and deposition of mercury into agriculture-associated water bodies.

Historically, Pacific salmon species have been an important source of sustenance for Indigenous Peoples in Northern Pacific latitudes^15^, including during pregnancy. However, a main concern of mercury contamination is exposure to methylmercury (MeHg), a component of total mercury that is neurotoxic and can cross the placenta to accumulate in the fetal brain^42^. This has important cultural and health implications for AI/AN Peoples that have relationships with Pacific salmon as a traditional food, as many individuals will consume Pacific salmon frequently. For example, among Peoples of the Columbia River Basin (Pacific Northwest, United States), a typical Tribal member consumes an average of 16 servings of Pacific salmon per month, with some Tribal members consuming nearly 60 servings per month (compared to an average of 3 servings per month in the general United States population)^43^.

There is a growing body of evidence supporting the conclusion that impacts of methyl mercury on fetal and infant development are far outweighed by the high nutritional benefits of consuming fish and other seafood (e.g., PUFAs). For example, results from the Seychelles Child Development Study, which studied the effects of MeHg on fetal neurological development from mothers in the Republic of Seychelles found no consistent adverse effects of MeHg from high seafood consumption on offspring neurological development at 22 and 24 years of age and with 10 total evaluations through infancy to adulthood^44^. However, consensus has yet to be reached regarding new evidence in fetal neurodevelopment and maternal MeHg exposure. MeHg was not specifically measured in these samples, but models for MeHg exposure during fish consumption by Li et al. (2024)^45^ showed lower MeHg concentrations in seafood from higher latitudes (compared to tropical latitudes), and that subsistence communities within United States territories (as an aggregated estimate of all subsistence communities) may consume >170g of seafood per day before exceeding MeHg EPA safe reference doses for chronic oral exposure (0.1 μg per kg BW per day^46^). It is unclear if levels of mercury measured in coho Pacific salmon here are of concern, but these estimates suggest that coho Pacific salmon are safe and beneficial at frequent human consumptions.

Effect sizes of our best performing model describing the occurrence of AI/AN pregnancy-related maternal death (Model J) indicate that Native food security in a multivariate context is a main variable informing this occurrence. Although the specific USDA food access research atlas variables used in the Native food security impacts variable will also impact Western food security, we did not seek to draw contrasts between roles of Native and Western food security in AI/AN pregnancy health. Rather, we aimed to explore how known factors affecting access to (and achievement of) Native food security may be related to AI/AN maternal health outcome.

The appropriateness of this approach is supported by the absence of multicollinearity between the Native food security impacts variable and the Western food insecurity experienced by the general population of a county. The reduction in coho Pacific salmon fatty acids that are beneficial to pregnancy, as well as the increases in total mercury, associated with increases in agricultural intensity support the need to consider quality of traditional foods when present (in addition to availability). Indeed, inadequate food quality rather than food quantity was associated with increased prevalences of obesity, diabetes, and hypertension in an AI/AN study group living within Chickasaw and Choctaw Nation territories^8^.

This work is consistent with concepts present within many traditional and cultural knowledge systems which link food, culture, and health. This is exemplified in a sentiment from an interviewed Indigenous leader, Sonia Quispe (Quechua Peoples, Rosaspata community, Peruvian Andes), who expressed sadness when traditional foods did not grow because it signaled to her that *Pachamama* (Quechua name for the sacred space where all human and non-human relatives among land, water, animals, and plants exist) was unwell^47^. Understanding that these links are present contributes to concepts that operationalize sustainability of food systems within traditional ecological knowledge bases via reciprocal relationships with environments (i.e., care for the land in perpetuity means the land cares for Peoples in perpetuity)^47^. It is understood within many Indigenous communities that Native food security depends on I/TFS^8,13,14,48^, in part because it recognizes that traditional foods may not persist without Indigenous self-determination and the full participation of Indigenous Peoples in their cultural and religious relationships with the land, water, and their plant and animal relatives therein^8,13,14^. Importantly, I/TFS is compatible with goals held broadly across various medical, health, and political agencies for improving Indigenous health disparities because, through a Western lens, I/TFS is analogous to a wrap-around framework that addresses the physical, spiritual, relational, and mental wellbeing of Indigenous Peoples by emphasizing access to, and relationships with, nutritionally healthy, culturally appropriate traditional foods, and connections to land and other elements held in sacred relationship.

### 4.1 Limitations

A critical limitation of this work is that it does not explicitly consider impacts of AI/AN community-driven food (and particularly, I/TFS food sovereignty) programs that may have been active within the counties analyzed. Tribally operated hatcheries that propagate culturally important fish species like Pacific salmon are prominent examples. Others include traditional knowledge dissemination classes held by the Spokane Tribal Network’s Tribal Food Sovereignty initiative^20^; traditional foods preparation and distribution to Alaska Natives carried out by the Native Conservancy’s Indigenous Food Sovereignty program^49^; and bison restoration efforts across Northern Great Plains territories supported by the Intertribal Buffalo Council^50^. Programs like these are only marginally considered in our analysis via the inclusion of AI/AN population counts, which makes significant assumptions about what the presence of AI/AN individuals, who may not be Indigenous to territories bounded within the specific counties, means for (Native) food security and AI/AN maternal health outcome. Considering the overarching relationships of Native food security to AI/AN maternal health detected in this study, we hypothesize that relationships between Native food security and maternal health will strengthen when these programs are appropriately integrated into future analyses.

Other limitations are that maternal characteristics were not considered. This was a consequence of study design, as our aim was to investigate broad patterns of food security and land use that may inform AI/AN pregnancy-related maternal death occurrence rather than roles for maternal characteristics that interact with a lived environment. Also, specific AI/AN Tribes and Nations which individuals belonged to, as well as resolution finer than county-level, were not available. However, it is expected that factors influencing maternal health will change in severity and type across communities and geographies. Maternal characteristics such as maternal age, race, BMI, parity, medical insurance status, and others can have profound impacts on maternal health and pregnancy outcome as well. Future work would benefit from stratifying population risk based on interactions between maternal characteristics and county-level features.

We were also not able to account for all variables that may affect relationships identified in this work. For example, several facilitators and barriers to Native food security could not be represented by USDA food access research atlas variables, such as state and federal land use policies^17,18^, or the presence of partnerships between state and/or private entities and AI/AN-led groups that work to support Native food security^17^. Additionally, all confounders of coho salmon nutrition were not measured (e.g., fish sexual maturity), and specific agricultural activities were not compared (e.g., organic farms, specific crops). Therefore, the current study does not draw causative links or directionality between land use, Native and/or Western food securities, AI/AN maternal health outcomes, and traditional food nutritional quality.

## 5. Conclusions

We integrate established bodies of literature concerning ways that land use impacts both the natural environment and marginalized groups, with cultural and traditional knowledge systems that assert the interconnectedness of the natural environment with AI/AN health and cultural wellbeing. This research also addresses areas of critical concern: AI/AN pregnancy-related maternal death, and vital traditional foods. Our analyses of AI/AN pregnancy-related maternal death and coho Pacific salmon nutrition demonstrate that relationships exist between food security, land use, and AI/AN maternal health. This work provides support for the importance of programs that specifically promote Native (versus Western) food security. Furthermore, we identify a threshold of Native food security that may improve AI/AN maternal health outcomes which has potential health policy relevance. Future analysis of causality and direction of these relationships may be impactful for improving AI/AN maternal health disparities.

## Data Availability

Code and data for the analysis of coho Pacific salmon nutrition is available on GitHub (https://github.com/contessaricci/landuse-foodsecurity-AIANmatmort) Human data were obtained from the CDC Multiple Cause Mortality restricted use files are are available upon submission of application to the CDC https://github.com/contessaricci/landuse-foodsecurity-AIANmatmort

## Acknowledgements

We thank the fishers and businesses that provided Pacific salmon samples: Robert Roose at Stillaguamish Tribe of Indians Natural Resources, Jamie Gonzalez at Pacific Harvest, Aaron Brookes at the Jamestown S’Klallam Natural Resources Department Fisheries Division, Tony Forsman at Suquamish Seafoods, Tara Livingood-Schott at the Confederated Tribes of the Chehalis Reservation Natural Resources Department, Gregory Wolfe at the Yakama Nation Fisheries Mid-Columbia Coho Restoration Project, Craig Smith at the Nisqually Natural Resources Department Harvest Management Program, TK at Kelly’s Fresh Fish, Scott Steltzner at the Squaxin Island Tribe Natural Resources Department, and Tad Iritani at Washington State University; we thank Mohammed R. Islam, Ph.D at Palouse Environmental Services for conducting the nutritional analysis; we thank our colleagues who volunteered their time to help process Pacific salmon samples: Shubhanker Sircar, Sabrina Haney, Bradford Dimos, Evan Barnes, Max Butensky, Tait Algayer, Tholen Blasko, Alyssa Williams; we thank Blake A. Foraker at the Washington State University Meat Lab for donating facility space and resources to process and store Pacific salmon samples.

## References

1. Hoyert DL. Maternal Mortality Rates in the United States, 2019. 2021. Accessed 17 Jun 2025. 10.15620/cdc:103855

2. Samuels-Kalow ME, Agrawal P, Rodriguez G, et al. Post-Roe emergency medicine: Policy, clinical, training, and individual implications for emergency clinicians. Acad Emerg Med. Dec 2022;29(12):1414–1421. doi:10.1111/acem.14609

3. Fleszar LG, Bryant AS, Johnson CO, et al. Trends in State-Level Maternal Mortality by Racial and Ethnic Group in the United States. Jama. Jul 3 2023;330(1):52–61. doi:10.1001/jama.2023.9043

4. Trost S, Beauregard J, Chandra G, et al. Pregnancy-related deaths among American Indian or Alaska Native persons: Data from maternal mortality review committees in 36 US States, 2017–2019. 2022.

5. Trost S, Beauregard J, Chandra G, et al. Pregnancy-related deaths: data from maternal mortality review committees in 36 US states, 2017–2019. Education. 2022;45(10):1–0.

6. Gregory ECW, Ely DM. *Trends and Characteristics in Prepregnancy Diabetes: United States, 2016–*2021 Vol. 72. 2023. National Vital Statistics Reports. Accessed 22 Jun 2025. https://www.cdc.gov/nchs/data/nvsr/nvsr72/nvsr72-06.pdf

7. Gemmill A, Berger BO, Crane MA, Margerison CE. Mortality Rates Among U.S. Women of Reproductive Age, 1999–2019. American Journal of Preventive Medicine. 2022;62(4):548–557. doi:10.1016/j.amepre.2021.10.009

8. Blue Bird Jernigan V, Wetherill MS, Hearod J, et al. Food Insecurity and Chronic Diseases Among American Indians in Rural Oklahoma: The THRIVE Study. Am J Public Health. Mar 2017;107(3):441–446. doi:10.2105/ajph.2016.303605

9. Sanders MA, Oppezzo M, Skan J, Benowitz NL, Schnellbaecher M, Prochaska JJ. Demographic and cultural correlates of traditional eating among Alaska Native adults at risk for cardiovascular disease. PLoS One. 2022;17(9):e0275445. doi:10.1371/journal.pone.0275445

10. Puddephatt JA, Keenan GS, Fielden A, Reaves DL, Halford JCG, Hardman CA. ’Eating to survive’: A qualitative analysis of factors influencing food choice and eating behaviour in a food-insecure population. Appetite. Apr 1 2020;147:104547. doi:10.1016/j.appet.2019.104547

11. FAO. Rome Declaration on World Food Security and World Food Summit Plan of Action: World Food Summit, 13-17 November 1996, Rome, Italy. FOOD SECURITY DECLARATIONS PROGRAMMES OF ACTION 1996. United Nations; 1996.

12. Rabbitt MP, Hales LJ, Reed-Jones M. Food Security in the U.S. - Definitions of Food Security. United States Department of Agriculture (USDA). Accessed 18 Jun, 2025. https://www.ers.usda.gov/topics/food-nutrition-assistance/food-security-in-the-us/definitions-of-food-security

13. Maudrie TL, Nguyen CJ, Wilbur RE, et al. Food Security and Food Sovereignty: The Difference Between Surviving and Thriving. Health Promot Pract. Nov 2023;24(6):1075–1079. doi:10.1177/15248399231190366

14. Elm-Hill R, Webster RM, Allen A. Native Food Security, from Lack to Abundance Vol. 2. 2023. Distinguishing between Native Food Sovereignty and Native Food Security in Indian Country. Accessed 2 Feb 2025. https://www.firstnations.org/wp-content/uploads/2023/04/Native-Food-Security-04April2023.pdf

15. Ross JA. The Spokan Indians. Michael J. Ross; 2011.

16. Kuratko CN, Barrett EC, Nelson EB, Salem N, Jr. The relationship of docosahexaenoic acid (DHA) with learning and behavior in healthy children: a review. Nutrients. Jul 19 2013;5(7):2777–810. doi:10.3390/nu5072777

17. Grann A, Carlsson L, Mansfield-Brown K. Barriers and supports to traditional food access in Mi’kma’ki (Nova Scotia) Canadian Food Studies/La Revue Canadienne des études sure l’alimentation. 2023;10:65–85. doi:DOI: 10.15353/cfs-rcea.v10i1.571

18. Sowerwine J, Mucioki M, Sarna-Wojcicki D, Hillman L. Reframing food security by and for Native American communities: a case study among tribes in the Klamath River basin of Oregon and California. Food Security. 2019/06/01 2019;11(3):579-607. doi:10.1007/s12571-019-00925-y

19. Maudrie TL, Clyma KR, Nguyen CJ, et al. “It Matters Who Defines It” -Defining Nutrition through American Indian, Alaska Native, and Native Hawaiian Worldviews. Current Developments in Nutrition. 2024;doi:10.1016/j.cdnut.2024.104429

20. (STN) STN. Tribal Food Sovereignty. Spokane Tribal Network (STN). Accessed 3/15/2025, 2025. https://spokanetribalnetwork.org/tfs/

21. United States Department of Agriculture (USDA). 2022 *Census of Agriculture*. 2024. Accessed 3/15/2025. https://www.nass.usda.gov/Publications/AgCensus/2022/Full_Report/Volume_1,_Chapter_1_US/usv1.pdf

22. Centers for Disease Control and Prevention (CDC). Data from: Mortality Multiple Cause-of-Death restricted-use files (2016-2019, 2022). 2024. *Washington, D.C*.

23. United States Department of Agriculture (USDA). 2017 Census of Agriculture. https://www.nass.usda.gov/Publications/AgCensus/2017/

24. United States Department of Agriculture (USDA). 2022 Census of Agriculture. Accessed 12/5/2024. https://www.nass.usda.gov/Publications/AgCensus/2022/

25. Rhone A. Food Access Research Atlas. U.S. Department of Agriculture (USDA). Accessed 5 Sep 2024. https://www.ers.usda.gov/data-products/food-access-research-atlas/go-to-the-atlas

26. March of Dimes. Maternity Care Desert. Accessed 1/24/2025. https://www.marchofdimes.org/peristats/data?top=23

27. United States Census Bureau. Quick Facts. Accessed 12/5/2024. https://www.census.gov/quickfacts/

28. Centers for Disease Control and Prevention, Statistics NCfH. System, Natality on CDC WONDER Online Database. Data are from the Natality Records 2016-2023, as compiled from data provided by the 57 vital statistics jurisdictions through the Vital Statistics Cooperative Program Accessed 1/15/2025. https://wonder.cdc.gov/

29. QGIS Geographic Information System. Version 3.34. 2025. QGIS.org

30. R: A Language and Environment for Statistical Computing. R Foundation for Statistical Computing; 2024. https://www.R-project.org/

31. Pham-Gia T, Hung TL. The mean and median absolute deviations. Mathematical and Computer Modelling. 2001/10/01/ 2001;34(7):921-936. 10.1016/S0895-7177(01)00109-1

32. Chesnaye NC, Stel VS, Tripepi G, et al. An introduction to inverse probability of treatment weighting in observational research. Clin Kidney J. Jan 2022;15(1):14–20. doi:10.1093/ckj/sfab158

33. McCaffrey DF, Griffin BA, Almirall D, Slaughter ME, Ramchand R, Burgette LF. A tutorial on propensity score estimation for multiple treatments using generalized boosted models. Stat Med. Aug 30 2013;32(19):3388–414. doi:10.1002/sim.5753

34. Ashman AM, Brown LJ, Collins CE, Rollo ME, Rae KM. Factors Associated with Effective Nutrition Interventions for Pregnant Indigenous Women: A Systematic Review. J Acad Nutr Diet. Aug 2017;117(8):1222–1253.e2. doi:10.1016/j.jand.2017.03.012

35. Jernigan VBB, Huyser KR, Valdes J, Simonds VW. Food Insecurity among American Indians and Alaska Natives: A National Profile using the Current Population Survey-Food Security Supplement. J Hunger Environ Nutr. 2017;12(1):1–10. doi:10.1080/19320248.2016.1227750

36. Egeland GM, Johnson-Down L, Cao ZR, Sheikh N, Weiler H. Food insecurity and nutrition transition combine to affect nutrient intakes in Canadian arctic communities. J Nutr. Sep 2011;141(9):1746–53. doi:10.3945/jn.111.139006

37. Yuzicapi ELG, Fidji Bouch-van Dusen, Rosella. Dakota and Lakota Traditional Food and Tea: Teachings from Elder Lorraine Yuzicapi. Journal of Indigenous Wellbeing: Te Mauri – Pimatisiwin. 2013;11(1):33.

38. Miller KM, Schulze AD, Ginther N, et al. Salmon spawning migration: Metabolic shifts and environmental triggers. Comparative Biochemistry and Physiology Part D: Genomics and Proteomics. 2009/06/01/ 2009;4(2):75–89. 10.1016/j.cbd.2008.11.002

39. Pickhardt PC, Stepanova M, Fisher NS. Contrasting uptake routes and tissue distributions of inorganic and methylmercury in mosquitofish (Gambusia affinis) and redear sunfish (Lepomis microlophus). Environmental Toxicology and Chemistry. 2006;25(8):2132–2142. doi:10.1897/05-595r.1

40. Stevenson LM, Matson PG, Pilla RM, et al. Analysis of biokinetic parameters reveals patterns in mercury accumulation across aquatic species. Science of The Total Environment. 2025/01/10/ 2025;959:178129. 10.1016/j.scitotenv.2024.178129

41. Ottesen RT, Birke M, Finne TE, et al. Mercury in European agricultural and grazing land soils. Applied Geochemistry. 2013/06/01/ 2013;33:1-12. 10.1016/j.apgeochem.2012.12.013

42. Li P, Feng X, Qiu G. Methylmercury Exposure and Health Effects from Rice and Fish Consumption: A Review. International Journal of Environmental Research and Public Health. 2010;7(6):2666–2691.

43. Schick T, Miller M. The U.S. Promised Tribes They Would Always Have Fish, but the Fish They Have Pose Toxic Risks. ProPublica. Accessed 3/15/2025. https://www.propublica.org/article/how-the-us-broke-promise-to-protect-fish-for-tribes

44. van Wijngaarden E, Thurston SW, Myers GJ, et al. Methyl mercury exposure and neurodevelopmental outcomes in the Seychelles Child Development Study Main cohort at age 22 and 24years. Neurotoxicol Teratol. Jan-Feb 2017;59:35–42. doi:10.1016/j.ntt.2016.10.011

45. Li M-L, Thackray CP, Lam VWY, Cheung WWL, Sunderland EM. Global fishing patterns amplify human exposures to methylmercury. Proceedings of the National Academy of Sciences. 2024;121(40):e2405898121. doi:doi:10.1073/pnas.2405898121

46. Joint FAO/WHO Expert Committee on Food Additives (JECFA). Methylmercury. Evaluations of the joint FAO/WHO. 2007. Accessed 3/15/2025. https://apps.who.int/food-additives-contaminants-jecfa-database/Home/Chemical/3083

47. Huambachano MA. Indigenous food sovereignty Reclaiming food as sacred medicine in Aotearoa New Zealand and Peru. New Zealand Journal of Ecology. 2019;43(3):1–6.

48. Blue Sky Minds Nonprofit, Washington State University, Council ORF. Okanogan Region Local Community Food System Assessment. 2024. Accessed 5/7/2025. https://static1.squarespace.com/static/642492ad34f4c9023a6143df/t/66ec8c514943b56429e62761/1726778452438/Okanogan-AssessmentBook_Sept2024_SinglePgs.pdf

49. Conservancy N. Indigenous Food Sovereignty. Native Conservancy. Accessed 3/15/2025, 2025. https://www.nativeconservancy.org/indigenous-food-sovereignty.html

50. Intertribal Buffalo Council. FOOD SOVEREIGNTY THROUGH BUFFALO RESTORATION. Intertribal Buffalo Council. Accessed 3/15/2025, 2025. https://www.itbcbuffalonation.org/copy-of-herd-development-grant-1

