## supplementary_material for "Land Use Barriers to Native and Western Food Security are Associated with Indigenous Maternal Death Occurrence and Changes to Nutritional Value of a Key Traditional Food"

### SUPPLEMENTARY METHODS

*Author positionalities*

CR is a cis Filipina and Italian woman who is an academic researcher at a land grant university (Washington State University) and is funded as an NIH fellow at time of publication (NIH 5F32MD019202). AO is cis Taiwanese American woman who is attending the University of British Columbia for her doctorate. She is studying Indigenous Resource Management in agroecological wetlands. MW is a citizen of the Spokane Tribe of Indians, an Interior Salish speaking people located on the Upper Columbia Plateau of what is currently called the USA. MW has a doctorate from the University of Hawaiʻi at Mānoa in Community and Cultural Concentration in Psychology and currently works with the Tribal Food Sovereignty and Indigenous Birth Justice teams at Spokane Tribal Network, a small non-profit on the Spokane Indian Reservation. LP is a cis woman who descends from the Spokane Tribe of Indians as well as German/Irish/English settlers and mother to a Hispanic (Mexican)/African American/Caucasian/Native American son. LP is a chef and currently works on the Tribal Food Sovereignty and Indigenous Birth Justice projects at the Spokane Tribal Network with her mother (MW). PH is a graduate of Washington State University currently working in aquaculture, providing health care to salmonid species for global food demand. AL is a Latino man who is a fish biologist for the Bureau of Reclamation in Yakima, Washington. AL is also a Ph.D. candidate at Washington State University in the School of Biological Sciences. MP is an Assistant Professor researching salmon physiology, conservation, and aquaculture. JP is a cisgender white woman who is a nurse and academic researcher at a land grant university (Washington State University). LH is a cisgender white women who is a public health scientist and academic researcher at a land grant university (Washington State University).

*CDC maternal death data*

Data years 2020 and 2021 were excluded due to changes in medical and maternal care that arose during the COVID-19 pandemic (e.g., complications exacerbated by hospital bed availability). Data years post-2022 were excluded due to changes in medical and maternal care resulting from the 2023 *Dobbs Decision* (e.g., geographic differences for receiving life-saving maternal emergency care[1,2]).

State and county associated with mortality records reflect state and county of residence. Records citing causes of death classified as vehicular, neoplasm, COVID-19 or respiratory infection (2022 records only), homicide, and exposure were excluded (n = 94). Records citing mental health-related causes (suicide, substance use) were included (n = 49) because there is are established relationship between pregnancy, mental health, and diet[3]. Robustness analysis[4,5] between inclusion and exclusion of records citing mental health-related causes of death were conducted to confirm that inclusion of these records did not meaningfully alter conclusions.

*USDA agricultural census data*

AI/AN pregnancy-related maternal deaths occurring in data years 2016-2019 were associated with agriculture acreage estimates in the 2017 agricultural census and deaths occurring in data year 2022 were associated with estimates in the 2022 agricultural census. Year of agricultural census used by non-observation counties was the 2017 agricultural census by default. For states where AI/AN pregnancy-related maternal deaths occurred in 2022, a random subset of non-observation counties was assigned the 2022 agricultural census that was equal to the total number of AI/AN pregnancy-related maternal deaths in the state which occurred in 2022. Year of agricultural census used for non-observation counties was also the year assigned for model weighting.

### Supplementary material

*USDA food access research atlas (FARA)*

The 2019 USDA FARA contains 148 variables relating to Western food security down to census tract resolution and is an update to the 2015 USDA FARA survey. We used the count from county AI/AN population determined to be at low food access (e.g., grocery stores, supermarkets) at ≥0.5 mi for county AI/AN-specific Western food insecurity, and we used the count of county general population determined to be at low income and low food access at ≥0.5 mi for county generalized Western food insecurity (supplementary table 1). Distance and economic barriers to food access are established proxies for quantifying food security in academic literature[6,7] and for public policy purposes[8]. We apply >0.5 mi regardless of urban and rural residence because it has been shown that this can more accurately describe rates of Western food insecurity[6].It has also been shown that including vehicle insecurity as a criterion for Western food insecurity improves accuracy of Western food insecurity estimates[6], but we did not include vehicle insecurity in our definition. This was because vehicle insecurity was included in our summary statistic for Native food security impacts. While there are many studies that also exclude vehicle insecurity in Western food insecurity estimates, we acknowledge this as a limitation.

Our Native food security impacts variable collapsed urbanization (proportion of census tracts designated as urban), median family income (averaged median family income across census tracts), AI/AN population count (sum of AI/AN population across census tracts), and vehicle insecurity (proportion of households across census tracts that do not have vehicle access)(supplementary table 1).

*GIS*

The 15 mi radius for our buffer zones was selected to account for the highly mobile nature of coho Pacific salmon during spawning migrations to breeding grounds, while also avoiding high overlap of catch location buffer zones. Climate and agricultural intensity values were calculated using the zonal statistics plugin[9], with the respective variable map as the raster layer and the buffer zones as the polygon layer.

Temperature and precipitation measurements were averaged for the month prior to catch date to account for sampling bias (e.g., collecting during favorable weather conditions) as well as recent accumulated climate conditions that may influence nutritional characteristics. Because temperature and precipitation maps were by default averaged over calendar month (versus a rolling average of 24h days), samples collected in the first half of a month were associated with average temperature and precipitation values for the month previous to catch, while sample collected in the second half of a month were associated with values for the concurrent month of catch.

Temperature was the month average of daytime temperatures (degrees Celsius, °C) estimated by the month Land Surface Temperature map from NASA Earth Observations (NEO)[10]. Precipitation was the month average of the 24 h daily precipitation total (inches, in) estimated using the National Weather Service month to date maps[11]. Urbanization was estimated using the Multi-Resolution Land Characteristics (MRLC) consortium 2019 Percent Developed Imperviousness map[12]. Agricultural intensity was estimated using the MRLC National Land Cover Database map[13] and the sum of pixels classified as "Plant/cultivated" (i.e, pasture/Hay, cultivated crops).

*Sampling*

All samples were obtained from Indigenous fishers or Indigenous-owned businesses except for Clearwater River samples, which were obtained from a non-Indigenous local fisher. Fishers and businesses provided 4 fish technical replicates per catch location (2 male, 2 female fish). Coho

### Supplementary material

Pacific salmon samples were obtained from: Stillaguamish Tribe of Indians (n=4), Muckleshoot Indian Tribe (n=4), Jamestown S'Klallam Tribe (n=4), Confederated Tribes of the Chehalis (n=4), Yakama Nation (n=4), Nisqually Indian Tribe (n=8), Squaxin Island Tribe (n=4), and Suquamish Tribe (n=4). We additionally obtained fish from an Indigenous owned (Tsm'syen First Nations) business caught in Makah Tribe territory (n=4), and fish from a local, non-Indigenous fisher caught in Nez Perce Tribe territory. All fishers and businesses were compensated at asking price, with some choosing to donate samples for the study. All fishers and businesses recorded the sex of the fish and GPS coordinates of catch locations. Some fishers or businesses provided whole fish while others chose to provide a fillet only. If fishers or businesses provided a fillet only, the fork length, presence/absence of a hatchery mark (fin clip), and 5-10 scales were provided by the fishers. Fork length was used to infer fish weight in kg[14] while scales were used to estimate fish age in yrs[14]. If the fishers or businesses chose to provide whole fish, the information and scale samples were collected in house. Because our sampling design relied on fishers and businesses rather than researcher field sampling, a standardized estimation for fish sexual maturity could not be obtained and we recognize this as a limitation. Samples that were shipped were shipped frozen overnight. All samples were stored long term at -80 °C until muscle tissue collection for nutritional analyses.

Although all samples at all geographic locations were caught while fillet quality was high and were also representative of fish that are consumed in AI/AN and other communities, sexual maturation can influence Pacific salmon nutritional status[15,16] and we acknowledge this as a limitation of our sampling strategy. All fish samples were frozen at -20 °C on day of catch.

*Nutritional analysis*

Because our sampling strategy did not allow collection of muscle tissue upon catch and prior to freezing, all fish were first thawed at 4 °C overnight before muscle tissue collection. Additionally, measuring nutritional profiles from fish at this stage is an accurate reflection of the nutritional status of coho Pacific salmon when AI/AN and other communities would consume these fish. After collection, muscle tissues were re-frozen at -80 °C and shipped overnight to Palouse Environmental Services labs. Proximate variables were reported as weight per 100 g of tissue. FAMEs were reported as weight per crude fat per 100 g of tissue. THg was reported as parts per million (ppm). We do not assess effects of heterogeneity in freezing and storage methods on nutritional analysis[17] and we recognize this as a limitation of the work.

*Statistical analysis*

The MAD cutoff for our analysis of AI/AN pregnancy-related maternal death occurrence was relaxed to 4 standard deviations. We applied the outlier identification cutoff at 4 standard deviations because applying the common practice of 3 standard deviations heavily skewed outlier identification toward mortality observations (>76% of total outliers identified at 3 standard deviations), which reduced the mortality sample size by approximately 10% (versus 3% for the no-observation sample size).

Weak to moderate multicollinearity was detected between independent variables included in AI/AN pregnancy-related maternal death models but was within acceptable margins[18] (supplementary figure 2). Because the correlation between generalized Western food insecurity and AI/AN-specific Western food insecurity was high (Spearman's $\rho > 0.6$, supplementary figure 2) we did not include generalized Western food insecurity and AI/AN-specific Western food insecurity in the same model despite this still being within an acceptable level of multicollinearity[18]. Urbanization was initially considered for coho Pacific salmon nutritional analyses and defined as % impermeable surface within the 15 mi buffer zone. However, due to moderate multicollinearity between urbanization and climate variables (supplementary figure 3) which increased after calculating our “climate” variable (supplementary figure 4), urbanization was excluded from coho Pacific salmon nutritional analyses.

Weighted models for our analysis of AI/AN pregnancy-related maternal death were developed using a forward stepwise strategy to test mortality relationships in univariate models (Native food security impacts, generalized Western food insecurity, AI/AN-specific Western food insecurity, acreage dedicated to large agricultural acreage) and then multivariate models thereafter (supplementary table 3). Differences in scale of model variables were addressed by applying the robust scaler[19].Goodness of fit was calculated using the Performance package[20] in R. Redundancy analysis (RDA) was conducted using the vegan package[21] and the ggordiplots package[22] in R.

### SUPPLEMENTARY RESULTS

*Robustness analysis*

In our robustness analysis, non-observation counties were randomly removed to equal the total number of observation counties where mortalities did not result from mental health-related causes. Statistical protocols were carried out on this reduced dataset to determine if this change to the inclusion/exclusion impacted conclusions drawn (supplementary table 5).

Model J was still the most informative model in robustness analysis (supplementary table 5), but only Native food security impacts reached statistical significance under the Bonferroni multiple comparisons significance threshold (supplementary table 6). However, that this model remained the most informative by several ranks suggests that the explanatory power of the Native food security impacts variable relies on a multivariate context.

### SUPPLEMENTARY TABLES AND FIGURES

**Supplementary table 1 USDA FARA variables.** Variables included in the 2019 USDA FARA survey that were used to calculate variables in analysis of AI/AN pregnancy-related maternal death occurrence. Table shows the main model component, the specific analytical variable, and the calculation using USDA FARA variables.

| Model component | Analytical variable | Analytical variable calculation |
|---|---|---|
| Western food insecurity | AI/AN-specific Western food insecurity | Sum **laaianhalf** across county census tracts ÷ Sum **POP2010** across county census tracts |
| | Generalized Western food insecurity | Sum **lalowihalf** across county census tracts ÷ Sum **POP2010** across county census tracts |
| Native food security impacts | County urbanization | Sum **Urban** across county census tracts ÷ Total county census tracts |
| | Generalized low vehicle access | Sum **TractHUNV** across county census tracts ÷ Sum **OHU2010** across county census tracts |
| | County median family income | Sum **MedianFamilyIncome** across county census tracts ÷ Total county census tracts |
| | AI/AN population | Sum **TractAIAN** across county census tracts |

*Bold variables are names of specific variables included in 2019 USDA FARA that were used*

**Supplementary table 2.** Barriers and facilitators to traditional food access from academic literature sources used to inform the “Native food security impacts” variable. Corresponding 2019 USDA FARA variables that were used in PCA are shown.

| Barriers | FARA variable |
| --- | --- |
| Identity loss[a,b]<br>Knowledge loss[a,b,c,d]<br>Limited community support[a,b,d]<br>Lack of community programs[a,b] | *Total AI/AN population* |
| Costs of accessing traditional foods[a,b,c]<br>Cost of accessing knowledge[a] | *Median income* |
| Owning a vehicle[b,d] | *Vehicle insecure designation* |
| Urban residence[b,f] | *Urban designation* |
| Private land ownership[a,b]<br>Federal resource/land policies[a,b,d,f]<br>Land degradation[a,b,d,f]<br>Native foods contamination/quality[a,b,c,d,f]<br>Climate change[b,d,f]<br>Not enough Native foods[d,f] | |
| **Facilitators** | |
| Strength of cultural values[a,b,f]<br>Community engagement and activism[a,f]<br>Food sharing[b,f]<br>Sharing knowledge[b,f] | *Total AI/AN population* |
| External consultations and partnerships[a,f] | |

*Shaded cells are barrier/facilitator themes which can be represented by variables included in the USDA Food Access Research Atlas (FARA). Non-shaded cells are those which were not represented in “Native food security impacts” variable. Superscript letters correspond to referencing citing the barrier/facilitator.*

References for supplementary table 2:

a. Grann A, Carlsson L, Mansfield-Brown K. Barriers and supports to traditional food access in Mi’kma’ki (Nova Scotia) Canadian Food Studies/La Revue Canadienne des études sure l’alimentation. 2023;10:65-85. doi:DOI: 10.15353/cfs-rcea.v10i1.571

b. Sowerwine J, Mucioki M, Sarna-Wojcicki D, Hillman L. Reframing food security by and for Native American communities: a case study among tribes in the Klamath River basin of Oregon and California. *Food Security*. 2019/06/01 2019;11(3):579-607. doi:10.1007/s12571-019-00925-y

c. Batal M, Chan HM, Fediuk K, et al. First Nations households living on-reserve experience food insecurity: prevalence and predictors among ninety-two First Nations communities across Canada. *Can J Public Health*. Jun 2021;112(Suppl 1):52-63. doi:10.17269/s41997-021-00491-x

d. Sowerwine, Jennifer et al. “Enhancing Indigenous Food Sovereignty and Community Health Through the Karuk Agroecosystem Resilience Initiative: We Are Caring for It: xúus nu'éethti.” *Health promotion practice* vol. 24,6 (2023): 1096-1100. doi:10.1177/15248399231190368

e. Cidro J, Adekunle B, Peters E, Martens T. Beyond Food Security Understanding Access to Cultural Food for Urban Indigenous People in Winnipeg as Indigenous Food Sovereignty. Canadian Journal of Urban Research. 2015;24(1):24-43.

f. Blue Sky Minds Nonprofit, Washington State University, Council ORF. Okanogan Region Local Community Food System Assessment. 2024. Accessed 5/7/2025. https://static1.squarespace.com/static/642492ad34f4c9023a6143df/t/66ec8c514943b56429e62761/1726778452438/Okanogan-AssessmentBook_Sept2024_SinglePgs.pdf

**Supplementary table 3 Description of models.** Models compared in analysis of AI/AN pregnancy-related maternal death occurrence. Variables are aggregated at county level.

| Model Level | Model | Variables |
|---|---|---|
| Univariate | A | Generalized Western food insecurity |
| | B | AI/AN-specific Western food insecurity |
| | C | Native food security impacts |
| | D | Agricultural acreage |
| 2-variable multivariate | E | Native food security impacts * Generalized Western food insecurity |
| | F | Native food security impacts * AI/AN-specific Western food insecurity |
| | G | Generalized Western food insecurity * agricultural acreage |
| | H | AI/AN-specific Western food insecurity * agricultural acreage |
| | I | Native food security impacts * agricultural acreage |
| 3-variable multivariate | J | Native food security impacts * Generalized Western food insecurity * agricultural acreage |
| | K | Native food security impacts * AI/AN-specific Western food insecurity * agricultural acreage |

*‘*’ indicates full-factorial designation for model definition*

**Supplementary table 4 Food secure counties with very high agricultural acreage**. Counts of non-observation vs. observation counties with very low Native food security impacts and/or high Western food security in association with very high agricultural acreage

| Very low Native food security impacts | | |
|---|---|---|
| Non-observation | Observation | Total |
| 8 | 3 | 11 |
| **High Western food security** | | |
| Non-observation | Observation | Total |
| 10 | 7 | 17 |
| **Very low Native food security impacts and high Western food security** | | |
| Non-observation | Observation | Total |
| 5 | 1 | 6 |

*Very high agricultural acreage: >3rd quartile (>845,289 acres); total counties with very high agricultural acreage: 51; non-observation counties with very high agricultural acreage: 24; observation counties with very high agricultural acreage: 27; very low Native food security impacts: <1st quartile (<-2.311); high Western food security: >median (>0. 287)*

**Supplementary Table 5 Robustness analysis model comparisons.** Model goodness of fit comparisons for county-level factors associated with AI/AN pregnancy-related maternal death occurrence. Models arranged in descending order from best fit to worst fit.

| Model | Variables | Tjur's $R^2$ | RMSE | PCP |
|---|---|---|---|---|
| RA_J | Native food, Western food (gen), ag acreage | 0.301 | 0.421 | 0.651 |
| RA_K | Native food, Western food (AI/AN), ag acreage | 0.293 | 0.428 | 0.647 |
| RA_E | Native food, Western food (gen) | 0.234 | 0.436 | 0.617 |
| RA_F | Native food, Western food (AI/AN) | 0.222 | 0.438 | 0.611 |
| RA_C | Native food | 0. 216 | 0.436 | 0.608 |
| RA_I | Native food, ag acreage | 0.213 | 0.442 | 0.607 |
| RA_H | Western food (AI/AN), ag acreage | 0.140 | 0.471 | 0.570 |
| RA_G | Western food (gen), ag acreage | 0.038 | 0.480 | 0.542 |
| RA_B | Western food (AI/AN) | 0.008 | 0.497 | 0.504 |
| RA_D | Ag acreage | 0.001 | 0.501 | 0.500 |
| RA_A | Western food (gen) | $3.298^{e-4}$ | 0.500 | 0.500 |

*Native food: county Native food security impacts; Western food (gen): county generalized Western food insecurity; ag acreage: county acreage dedicated to large agricultural operations; RMSE: root mean squared error; PCP: % correct predicted*

**Supplementary table 6 Model RA_J.** Results for the model most explanatory to AI/AN pregnancy-related maternal death occurrence in robustness analysis. Statistically significant effects were subject to a Bonferroni multiple comparisons threshold of p-value ≤ 0.004.

| | Est | SE | t val | p-val |
|---|---|---|---|---|
| **Native food** | **1.190** | **0.388** | **3.064** | **0.003** |
| Western food (gen) | 0.510 | 0.502 | 1.016 | 0.311 |
| Ag acreage | -0.318 | 0.407 | -0.782 | 0.436 |
| Native food X Western food (gen) | -0.539 | 0.526 | -1.026 | 0.307 |
| Western food (gen) X ag acreage | 0.375 | 0.553 | 0.678 | 0.499 |
| Native food X ag acreage | -0.381 | 0.305 | -1.247 | 0.214 |
| Native food X Western food (gen) X ag acreage | 0.473 | 0.395 | 1.196 | 0.234 |

*Native food: county Native food security impacts; Western food (gen): county generalized Western food insecurity; ag acreage: county acreage dedicated to large agricultural operations; Est: estimate; SE: standard error; ag: agriculture; null deviance: 394.73 on 149 degrees of freedom; residual deviance: 282.51 on 142 degrees of freedom; bolded p-values represent predictors meeting Bonferroni significance threshold*

**Supplementary table 7 Health benefits and risks of FAMEs.** Examples of known or putative health benefits and risks of fatty acid methyl esters (FAMEs) in coho Pacific salmon that were most responsive to agricultural intensity, with a focus on health benefits during pregnancy and to offspring.

| FAME | Health Benefit(s) | Reference |
|---|---|---|
| Docosahexaenoic Acid (DHA) | Infant brain development | Kuratko, Connye N et al. “The relationship of docosahexaenoic acid (DHA) with learning and behavior in healthy children: a review.” Nutrients vol. 5,7 2777-810. 19 Jul. 2013, doi:10.3390/nu5072777 |
| | Infant cognition | Judge MP, Harel O, Lammi-Keefe CJ. Maternal consumption of a docosahexaenoic acid–containing functional food during pregnancy: benefit for infant performance on problem-solving but not on recognition memory tasks at age 9 mo2. The American Journal of Clinical Nutrition. 2007/06/01/ 2007;85(6):1572-1577. doi:https://doi.org/10.1093/ajcn/85.6.1572 |
| Eicosadienoic Acid (EA) | Maternal gestational lipid metabolism | Li L-J, Lu R, Rawal S, et al. Maternal plasma phospholipid polyunsaturated fatty acids in early pregnancy and thyroid function throughout pregnancy: a longitudinal study. The American Journal of Clinical Nutrition. 2024/04/01/ 2024;119(4):1065-1074. doi:https://doi.org/10.1016/j.ajcnut.2024.02.016 |
| | Offspring lipid balance | Vidakovic AJ, Jaddoe VWV, Voortman T, Demmelmair H, Koletzko B, Gaillard R. Maternal plasma polyunsaturated fatty acid levels during pregnancy and childhood lipid and insulin levels. Nutrition, Metabolism and Cardiovascular Diseases. 2017/01/01/ 2017;27(1):78-85. doi:https://doi.org/10.1016/j.numecd.2016.10.001 |
| Gamma-Linolenic Acid (GLA) | Infant growth | Mychaleckyj JC, Zhang D, Nayak U, et al. Association of breast milk gamma-linolenic acid with infant anthropometric outcomes in urban, low-income Bangladeshi families: a prospective, birth cohort study. European Journal of Clinical Nutrition. 2020/05/01 2020;74(5):698-707. doi:10.1038/s41430-019-0498-6 |

| | | |
|---|---|---|
| | Infant cardiac development | Paredes A, Justo-Méndez R, Jiménez-Blasco D, et al. γ-Linolenic acid in maternal milk drives cardiac metabolic maturation. Nature. 2023/06/01 2023;618(7964):365-373. doi:10.1038/s41586-023-06068-7 |
| Myristic Acid (MA) | Infant appetite | Gutiérrez-García AG, Contreras CM, Díaz-Marte C. Myristic acid in amniotic fluid produces appetitive responses in human newborns. Early Human Development. 2017/12/01/ 2017;115:32-37. doi:https://doi.org/10.1016/j.earlhumdev.2017.08.009 |
| | Live birth | Kim K, Browne RW, Nobles CJ, et al. Associations Between Preconception Plasma Fatty Acids and Pregnancy Outcomes. Epidemiology. 2019;30 |
| Pentadecanoic Acid (PDA) | Infant growth | Ciesielski V, Guerbette T, Fret L, et al. Dietary pentadecanoic acid supplementation at weaning in essential fatty acid-deficient rats shed light on the new family of odd-chain n-8 PUFAs. The Journal of Nutritional Biochemistry. 2025/03/01/ 2025;137:109814. doi:https://doi.org/10.1016/j.jnutbio.2024.109814 |
| | Embryo development | Zarezadeh R, Nouri M, Hamdi K, Shaaker M, Mehdizadeh A, Darabi M. Fatty acids of follicular fluid phospholipids and triglycerides display distinct association with IVF outcomes. Reproductive BioMedicine Online. 2021/02/01/ 2021;42(2):301-309. doi:https://doi.org/10.1016/j.rbmo.2020.09.024 |
| Palmitoleic Acid (POA) | Offspring lipid metabolism | Niinistö S, Takkinen H-M, Uusitalo L, et al. Maternal dietary fatty acid intake during pregnancy and the risk of preclinical and clinical type 1 diabetes in the offspring. British Journal of Nutrition. 2014;111(5):895-903. doi:10.1017/S0007114513003073 |
| | Oocyte maturation | Mirabi P, Chaichi MJ, Esmaeilzadeh S, et al. The role of fatty acids on ICSI outcomes: a prospective cohort study. Lipids in Health and Disease. 2017/01/21 2017;16(1):18. doi:10.1186/s12944-016-0396-z |
| **Fame** | **Health Risk(s)** | **Reference** |
| Palmitic Acid (PA) | Placental inflammation | Shirasuna K, Takano H, Seno K, et al. Palmitic acid induces interleukin-1β secretion via NLRP3 inflammasomes and inflammatory responses through ROS production in human placental cells. Journal of Reproductive Immunology. 2016/08/01/ 2016;116:104-112. doi:https://doi.org/10.1016/j.jri.2016.06.001 |
| | Offspring heart disease | Zhao R, Cao L, Gu W-J, et al. Gestational palmitic acid suppresses embryonic GATA-binding protein 4 signaling and causes congenital heart disease. Cell Reports Medicine. 2023;4(3)doi:10.1016/j.xcrm.2023.100953 |

*Row colors delineate specific FAMEs and respective examples of health benefit(s) and risk(s)*

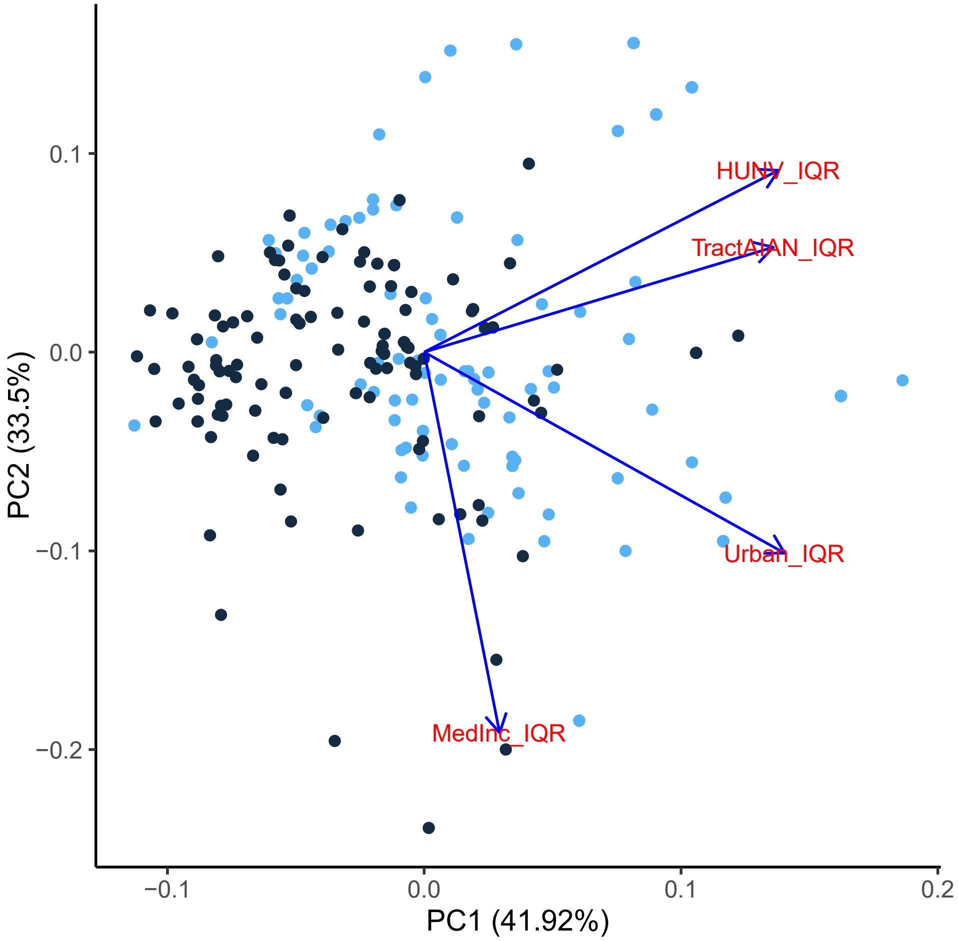


**Supplementary figure 1 "Native food security impacts" summary variable.** PC loadings for IQR normalized USDA FARA variables included in "Native food security impacts" summary variable. Blue arrows: eigenvectors for variables included in summary variable. Black dots: counties where mortality was not observed; Blue dots: counties where mortality was observed. HUNV_IQR: generalized vehicle insecurity; TractAIAN_IQR: county AI/AN population count; Urban_IQR: proportion of census tracts within county that are designated urban; MedInc_IQR: median family income averaged across county census tracts.

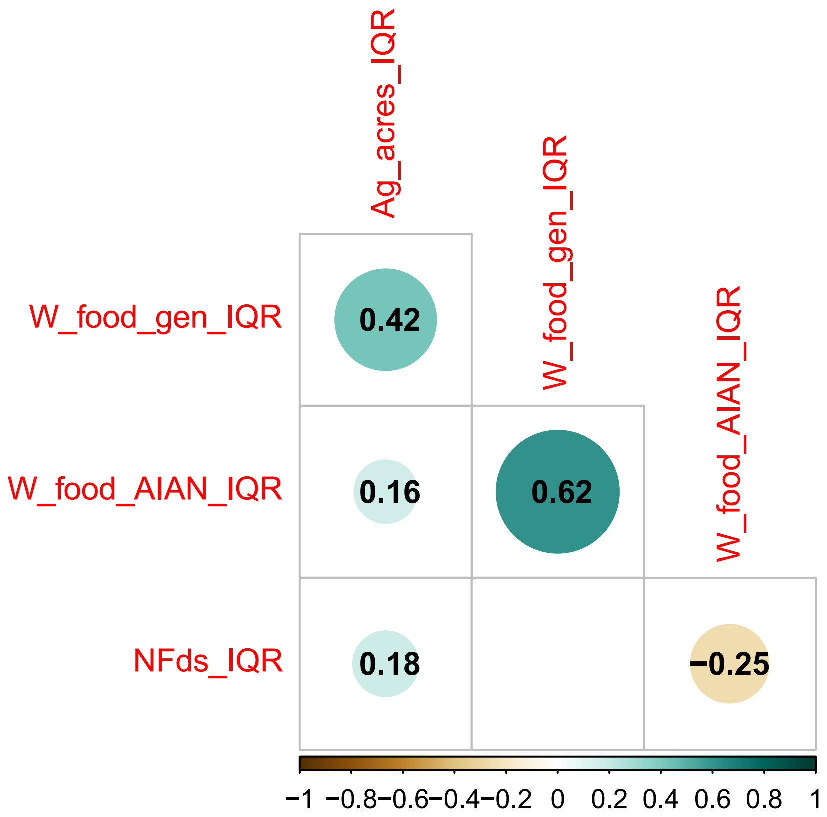


**Supplementary figure 2 Correlation plot for model components included in AI/AN pregnancy-related maternal death analysis.** Variables are IQR normalized. Size of circle represents strength of correlation, color of circle represents direction of correlation, black numbers represent correlation coefficient (Spearman's ρ). Ag_acres_IQR: acreage dedicated to large agricultural operations; W_food_gen_IQR: generalized Western food insecuirty; W_food_AIAN_IQR: AI/AN-specific Western food insecurity; NFds_IQR: “Native food security impacts” summary variable

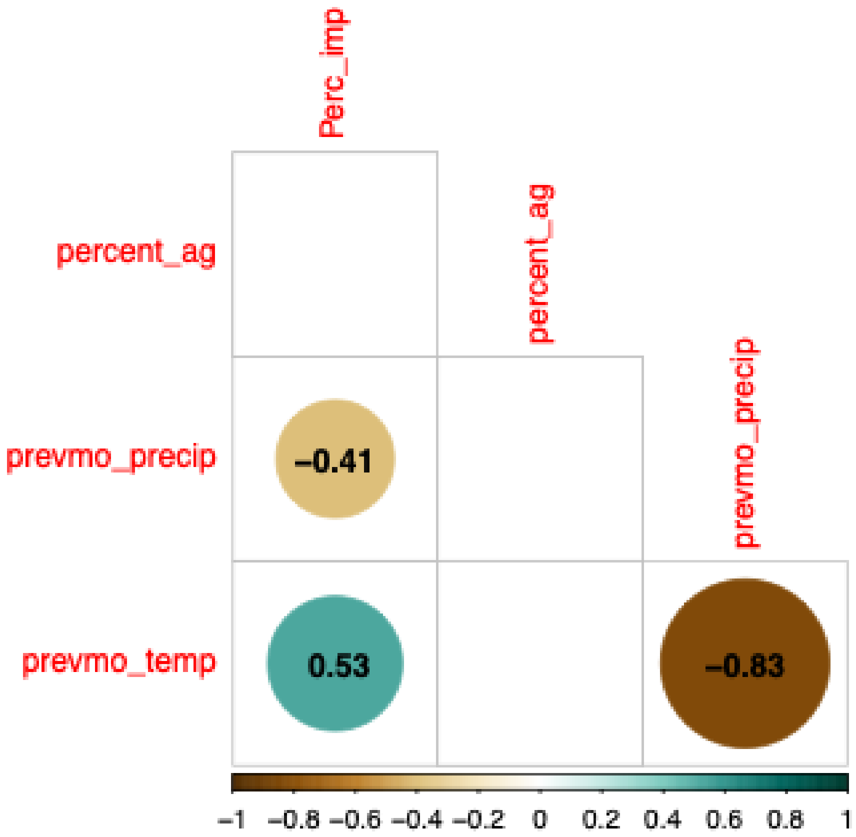


**Supplementary figure 3 Correlation plot for multicollinearity between coho Pacific salmon model components for nutritional analysis prior to creation of climate variable.** Size of circle represents strength of correlation, color of circle represents direction of correlation, black numbers represent correlation coefficient (Spearman's ρ). Perc_imp: % urbanization; percent_ag: % agriculture intensity; prevmo_precip: average precipitation (in) for month previous to catch date; prevmo_temp: average temperature (°C) for month previous to catch date.

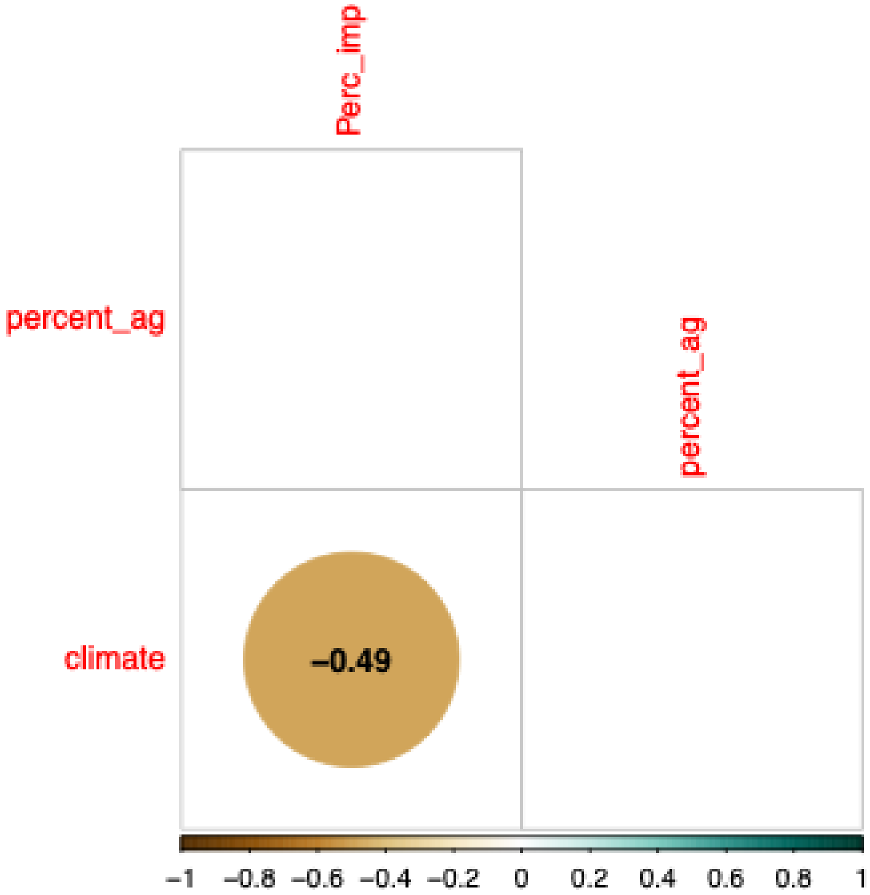


**Supplementary figure 4 Correlation plot for model components included in coho Pacific salmon nutritional analysis after creation of climate variable.** Size of circle represents strength of correlation, color of circle represents direction of correlation, black numbers represent correlation coefficient (Spearman's ρ). *Perc_imp: % urbanization;* percent_ag: % agriculture intensity; prevmo_precip: average precipitation (in) for month previous to catch date; prevmo_temp: average temperature (°C) for month previous to catch date.

**Supplementary material**

**References:**


1. Guttmacher Institute. Interactive Map: US Abortion Policies and Access After Roe. Accessed 4/15/2025. https://states.guttmacher.org/policies
2. Seitz A. Dozens of pregnant women, some bleeding or in labor, are turned away from ERs despite federal law. *The Associated Press*. Accessed 3/15/2025. https://apnews.com/article/pregnant-women-emergency-room-ectopic-er-edd66276d2f6c412c988051b618fb8f9
3. Trost S, Beauregard J, Chandra G, et al. Pregnancy-related deaths: data from maternal mortality review committees in 36 US states, 2017–2019. *Education*. 2022;45(10):1-0.
4. Rosenhead J. Robustness Analysis. *Wiley Encyclopedia of Operations Research and Management Science*. 2011.
5. Houkes W, Šešelja D, Vaesen K. Robustness Analysis. *The Routledge Handbook of Philosophy of Scientific Modeling*. 1st ed. Routledge; 2024:13:chap 14.
6. Salari M, Reyna M, Kramer MD, Taylor HA, Clifford GD. Food Desert Assessment: An Analytical Framework for Comparing Utility of Metrics and Indices; Case Study of Key Factors, Concurrences, and Divergences. *<h4 data-toggle="collapse" style="box-sizing: border-box; font-family: NexusSansWebPro; font-weight: 500; line-height: 26px; color: rgb(80, 80, 80); margin: 0px 0px 20px; font-size: 20px; cursor: auto; padding: 0px; caret-color: rgb(80, 80, 80);">SSRN*. 2021;doi:http://dx.doi.org/10.2139/ssrn.3823677
7. Ghosh-Dastidar B, Cohen D, Hunter G, et al. Distance to Store, Food Prices, and Obesity in Urban Food Deserts. *American Journal of Preventive Medicine*. 2014/11/01/ 2014;47(5):587-595. doi:https://doi.org/10.1016/j.amepre.2014.07.005
8. Ploeg MV, Dutko P, Breneman V. Measuring Food Access and Food Deserts for Policy Purposes. *Applied Economic Perspectives and Policy*. 2015;37(2):205-225.
9. *Zonal Statistics Plugin*. QGIS Association; 2024. https://github.com/qgis/QGIS/blob/master/INSTALL.md
10. NASA Earth Observations (NEO), Land Processes Distributed Active Archive Center (LPDAAC), Team MLS. LAND SURFACE TEMPERATURE [DAY] (1 MONTH - TERRA/MODIS)[map]. Moderate Resolution Imaging Spectroradiometer (MODIS). Greenbelt, Maryland: EOS Project Science Office; 2025. p. Map.
11. National Weather Service (NWS), (NOAA) NOaAA. Quantitative Precipitation Estimate (QPE) [map]. In: (NWS) NWPS, editor.: National Weather Service; 2023.
12. Dewitz J. National Land Cover Database (NLCD) 2019 Impervious Products [map]. U.S. Geological Survey data release; 2021.
13. MRLC Project. National Land Cover Database (NLCD)(CONUS) [map]. In: Multi-Resolution Land Characteristics (MRLC) Consortium, editor. Sioux Falls, SD: U.S. Geological Survey (USGS),
 Earth Resources Observation and Science (EROS) Center,
 MRLC Project; 2019.
14. Isely JJ, Grabowski TB. Age and Growth. In: Guy CS, Brown ML, eds. *Analysis and Interpretation of Freshwater Fisheries Data*. American Fisheries Society; 2007:187-228:chap 5.
15. Aussanasuwannakul A, Kenney PB, Weber GM, et al. Effect of sexual maturation on growth, fillet composition, and texture of female rainbow trout (Oncorhynchus mykiss) on a high nutritional plane. *Aquaculture*. 2011/07/04/ 2011;317(1):79-88. doi:https://doi.org/10.1016/j.aquaculture.2011.04.015
16. Bilinski E, Jonas REE, Peters MD, Choromanski EM. Effects of Sexual Maturation on the Quality of Coho Salmon (Oncorhynchus Kisutch) Flesh. *Canadian Institute of Food Science and Technology Journal*. 1984/10/01/ 1984;17(4):271-273. doi:https://doi.org/10.1016/S0315-5463(84)72570-3

17. Bao Y, Ertbjerg P, Estévez M, Yuan L, Gao R. Freezing of meat and aquatic food: Underlying mechanisms and implications on protein oxidation. *Compr Rev Food Sci Food Saf*. Nov 2021;20(6):5548-5569. doi:10.1111/1541-4337.12841
18. Gregorich M, Strohmaier S, Dunkler D, Heinze G. Regression with Highly Correlated Predictors: Variable Omission Is Not the Solution. *Int J Environ Res Public Health*. Apr 17 2021;18(8)doi:10.3390/ijerph18084259
19. de Amorim LBV, Cavalcanti GDC, Cruz RMO. The choice of scaling technique matters for classification performance. *Applied Soft Computing*. 2023/01/01/ 2023;133:109924. doi:https://doi.org/10.1016/j.asoc.2022.109924
20. Lüdecke D, Ben-Shachar MS, Patil I, Waggoner P, Makowski D. performance: An R Package for Assessment, Comparison and Testing of Statistical Models. *Journal of Open Source Software*. 2021;6(60):3139. doi:10.21105/joss.03139
21. *vegan: Community Ecology Package*. 2024. https://CRAN.R-project.org/package=vegan
22. *ggordiplots: Make 'ggplot2' Versions of Vegan's Ordiplots*. 2024. https://CRAN.R-project.org/package=ggordiplots
